# Demographic and genetic factors shape the epitope specificity of the human antibody repertoire against viruses

**DOI:** 10.1101/2023.11.07.23298153

**Authors:** Axel Olin, Anthony Jaquaniello, Maguelonne Roux, Ziyang Tan, Christian Pou, Florian Dubois, Bruno Charbit, Dang Liu, Emma Bloch, Emmanuel Clave, Itauá Leston Araujo, Antoine Toubert, Michael White, Maxime Rotival, Petter Brodin, Darragh Duffy, Lluis Quintana-Murci, Etienne Patin, Milieu Intérieur Consortium

## Abstract

Antibodies are central to immune defenses. Despite advances in understanding the mechanisms of antibody generation, a comprehensive model of how intrinsic and external factors shape human humoral responses to viruses is lacking. Here, we apply PhIP-Seq to investigate the effects of demographic and genetic factors on antibody reactivity to more than 97,000 viral peptides in 1,212 healthy adults. We demonstrate that age, sex, and continent of birth extensively influence the viruses and viral epitopes targeted by the human antibody repertoire. Among 108 lifestyle and health-related variables, smoking exerts the strongest, yet reversible, impact on antibody profiles, primarily against rhinoviruses. Additionally, we identify strong associations between antibodies against 34 viruses and genetic variants at *HLA*, *FUT2*, *IGH,* and *IGK* genes, some of which increase autoimmune disease risk. These findings offer a valuable resource for understanding the factors affecting antibody-mediated immunity, laying the groundwork for optimizing vaccine strategies.

---

Antibodies are essential effectors of humoral immunity and serve as correlates of protection following vaccination or natural infection. The cellular and molecular processes underlying antibody production and maintenance are thought to depend on diverse factors that collectively shape the strength and longevity of the antibody repertoire. Family- and population-based studies have uncovered marked differences in antibody titers with sex and age. For example, women often exhibit higher titers against human papillomavirus (HPV)^1^ and Epstein-Barr virus (EBV)^1,2^ and generally mount stronger vaccine responses than men^3^. Furthermore, antibodies against persistent herpesviruses like herpes simplex virus 1 (HSV-1) and cytomegalovirus (CMV) tend to increase with age, reflecting cumulative exposure^1,2,4,5^. In contrast, antibodies against viruses that primarily infect children (e.g., respiratory syncytial virus (RSV) and varicella-zoster virus (VZV)) or those included in immunization schedules (e.g., measles, mumps, and rubella viruses) typically persist at high levels in most adults^1,2^. Other non-genetic factors associated with antibody levels include socioeconomic status^1,2^ and smoking^4^.

Human genetic factors also affect antibody production. Total and virus-specific antibody titers against CMV, EBV, and influenza A virus (IAV) have been shown to be heritable^2,6–9^. At the genome-wide scale, the *HLA* locus presents strong associations with antibody titers against EBV, hepatitis B virus (HBV), VZV, and molluscum contagiosum virus^2,10–16^. Other loci, including *IGH, STING1,* and *FUT2*, have been associated with antibodies targeting IAV and norovirus^4,17^.

Despite advancements in characterizing the determinants of the antiviral antibody response, most studies have focused on a limited number of viruses, hindering a comprehensive understanding of humoral immunity across the broad spectrum of viruses infecting humans^18^. Furthermore, while antibodies targeting a single virus can recognize numerous epitopes – the portion of an antigen recognized by the immune system – variability in epitope reactivity among individuals infected with the same virus remains poorly understood. Factors such as ethnicity^19^ and age^20^ have been suggested to influence this variability, but the determinants of inter-individual differences in viral antigenic specificity are yet to be discovered.

In this study, we delineate the extent and drivers of variation in the epitope-specific antiviral antibody repertoire in humans using phage immunoprecipitation sequencing (PhIP-seq), a high-throughput method for assessing antibody-epitope interactions^21,22^. PhIP-seq has been used to characterize antibody repertoire changes across various diseases^5,23^ and to evaluate humoral immunity against bacteria and food allergens^4,24–26^. A virus-specific PhIP-seq implementation, VirScan^27^, which spans the complete peptidome of all known human viruses, has recently allowed to investigate the impact of measles infection on antibody profiles^28^, immune development in neonates^29^, and immunodominant epitopes^30,31^. Here, we applied the VirScan phage library to profile over 97,000 viral peptides in 1,212 healthy adults and integrated this information with comprehensive demographic, lifestyle, and genetic data. This approach enabled us to characterize differences in the viruses, viral proteins, and epitopes targeted by individual antibody profiles and to identify key factors shaping the natural breath and epitope specificity of the human antibody repertoire against viruses.

## Results

### Extensive diversity in the antiviral antibody repertoire of healthy adults

To assess the virome-wide antibody repertoire, we performed PhIP-seq on 900 plasma samples from the *Milieu Intérieur* (MI) cohort^32^, comprising individuals of European ancestry with a balanced distribution of sex and age (20-69 years; Fig. 1a). To validate findings from the MI cohort and explore population differences in humoral responses, we also applied PhIP-seq to 312 samples from the EvoImmunoPop (EIP) cohort^33^, comprising 100 and 212 Belgian residents born in either Central Africa or Europe, respectively, all male and aged 20 to 50 years (Fig. 1b). For both cohorts, we used the VirScan V3 library, encompassing 115,753 56-amino-acid-long peptide sequences^27^. After filtering for unique viral sequences, we obtained a final set of 97,978 peptides representing a wide range of viral families and species (Extended Data Fig. 1a,b). PhIP-seq read counts for each viral peptide were then converted into standardized *Z*-scores (Methods), which measure peptide-antibody interactions and have been shown to correlate strongly with antibody titers^27^.

**Fig. 1:**
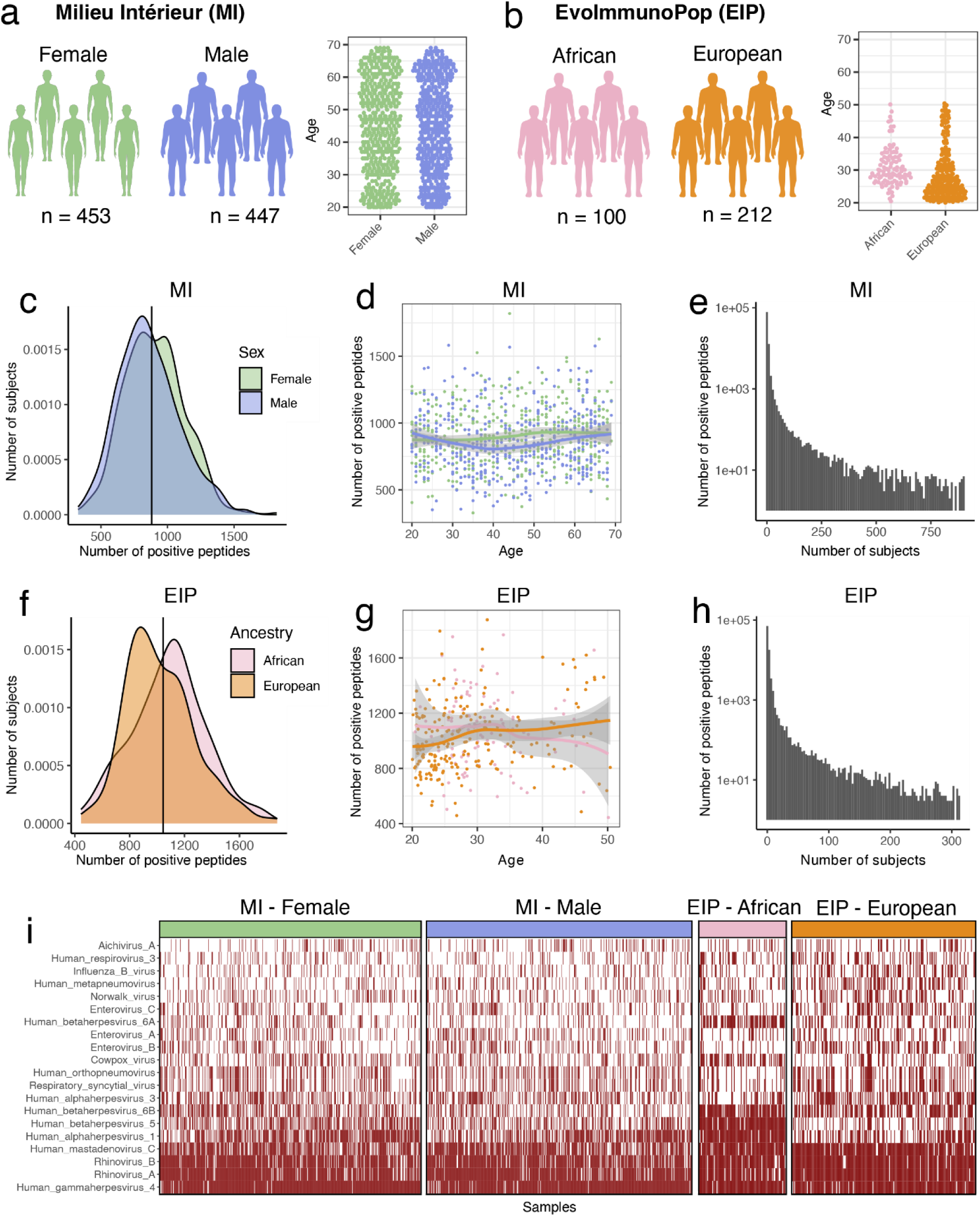
Assessing variation in the antibody repertoire in the Milieu Intérieur (MI) and EvoImmunoPop (EIP) cohorts. **a**, Sample sizes and age distribution by sex within the MI cohort. **b,** Sample sizes and age distribution by continent of birth within the EIP cohort. **c**, Density distributions of MI donors as a function of the number of peptides they react against, categorized by sex. **d**, Number of positive peptides per MI donor, as a function of age and sex. **e**, Number of peptides as a function of the number of positive MI donors. **f**, Density distributions of EIP donors as a function of the number of peptides they react against, categorized by continent of birth. **g**, Number of positive peptides per EIP donor, as a function of age and continent of birth. **h**, Number of peptides as a function of the number of positive EIP donors. **i**, Heatmap indicating the predicted infection status of each MI and EIP donor for the 20 most prevalent viruses, as determined by the AVARDA algorithm (*P*adj < 0.05 after Benjamini-Hochberg correction). The solid curves and shaded areas in **d** and **g** indicate the LOESS curves and the 95% confidence intervals.

The total number of positive peptides per individual was normally distributed (Fig. 1c,d,f,g), averaging 881 and 1,044 peptides for MI and EIP individuals, respectively, due to differences in cohort demographics, sampling protocols, or experimental batch effects (Methods).

Approximately 97% of peptides were positive in < 5% of individuals, reflecting individual-specific immunity (denoted *private* peptides) or false positives (Fig. 1e,h), consistent with previous reports^4,24,26^. As a result, we conducted all subsequent analyses on peptides positive in > 5% of individuals, with at least two peptides being positive from the same virus (denoted *public* peptides). In total, we identified 2,608 public peptides in MI and 3,210 in EIP, originating from 113 viral species, with EBV, IAV, and enterovirus B being the most prevalent in both cohorts (Extended Data Fig. 1c).

When investigating the reactivity of thousands of peptides simultaneously, the risk of cross-reactivity must be considered, as it can lead to false positives. To address this, we used the AVARDA algorithm, which estimates the probability of antibody reactivity per virus species, accounting for sequence alignment between peptides and library peptide representation^34^. As expected, seroprevalence determined by AVARDA was highest for common viruses such as EBV, HSV-1, CMV, rhinoviruses A and B, and adenovirus C in both cohorts (Fig. 1i). We validated the resolution, sensitivity, and serostatus prediction accuracy of both peptide-level *Z*-scores and virus-level AVARDA breadth scores through comparison with ELISA and Luminex assays (Methods; Supplementary Note; Supplementary Figs. 1 and 2; Table S1). Together, these analyses underscore the specificity and sensitivity of PhIP-seq results and reveal the extensive diversity of the human antibody repertoire targeting viruses causing common infections.

### Age and sex affect the breadth and epitope specificity of the antibody repertoire

Given the complementarity of peptide-level and AVARDA-based approaches (Supplementary Note), we explored the effects of non-genetic and genetic factors on antibody reactivity using peptide-level *Z*-scores and then verified whether the AVARDA breadth score for the corresponding virus was associated with the same factors. We first examined the effects of age and sex on the antiviral antibody repertoire, represented by the 2,608 public peptides and 132 AVARDA scores in the MI cohort. As no significant non-linear effects of age or age × sex interactions were observed, we only considered linear effects of age and sex (Methods). Linear regression modeling revealed that age is strongly associated with antibody reactivity against a broad range of viruses (Fig. 2a), in line with previous studies^4,5^.

**Fig. 2:**
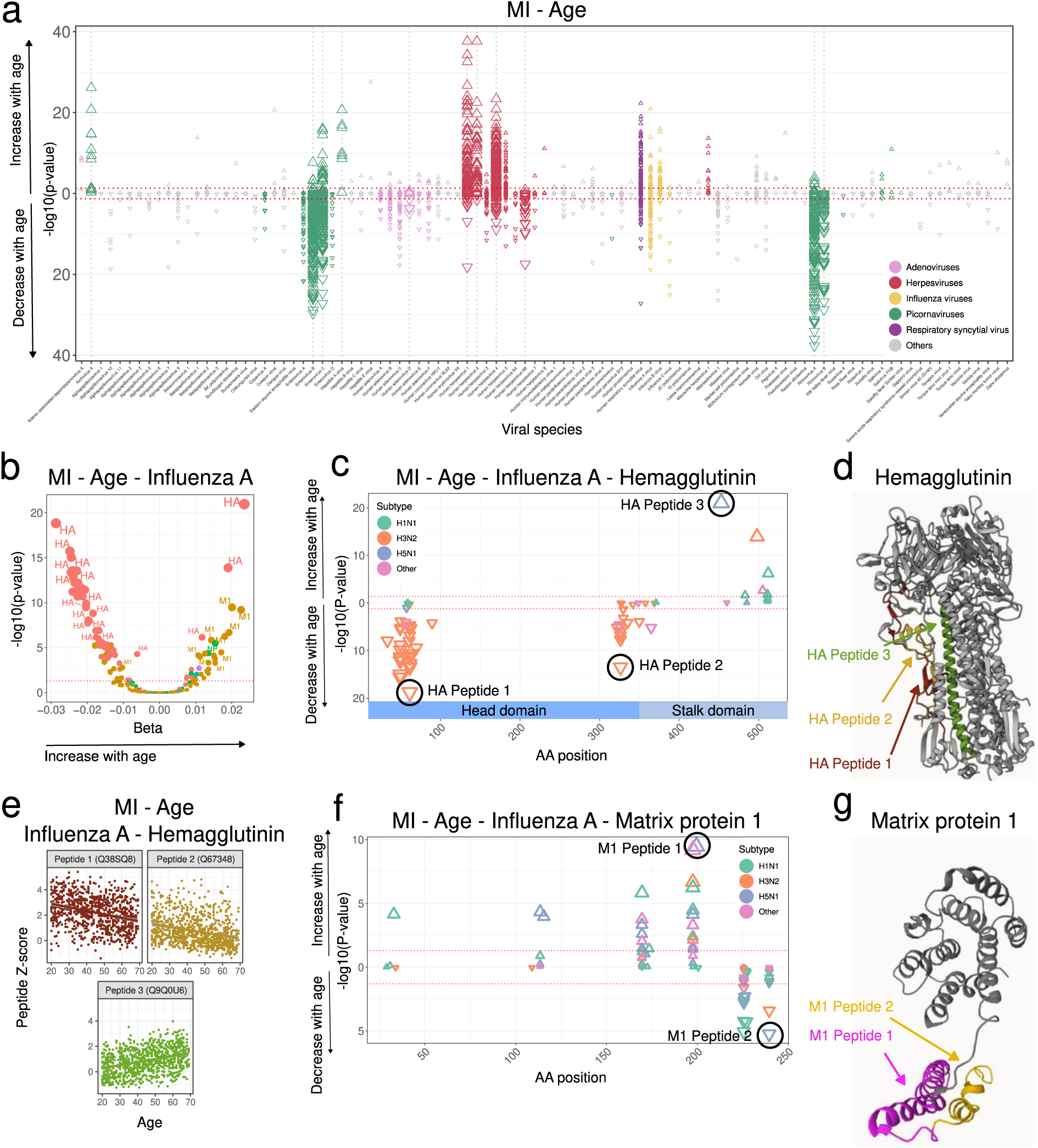
Age impacts the epitope-specific antiviral antibody repertoire. **a**, –log10(adjusted *P*-values) and direction of associations between all public peptide *Z*-scores and age in the MI cohort, by viral species. The dashed gray vertical lines indicate viruses for which the AVARDA breadth score is significantly associated with age. **b**, –log10(adjusted *P*-values) against effect sizes of associations between IAV peptide *Z*-scores and age in the MI cohort, colored by viral protein. **c**, Amino-acid positions of the midpoint of public HA peptides associated with age within the full IAV hemagglutinin (HA) protein for the MI cohort. The significance and direction of associations with age are indicated on the y-axis and by the direction of triangles, respectively. The triangle color indicates the IAV subtype. The most significant peptides for each epitope are indicated. **d**, Location of the peptides of interest indicated in (**c**) within the three-dimensional structure of HA. **e**, Antibody reactivity as a function of age for the HA peptides of interest highlighted in (**c**). **f**, Amino-acid positions of the midpoint of public M1 peptides associated with age within the full IAV Matrix Protein 1 (M1) protein for the MI cohort. The significance and direction of associations with age are indicated on the y-axis and by the direction of triangles, respectively. The triangle color indicates the IAV subtype. The most significant peptides for each epitope are indicated. **g**, Location of the peptides of interest indicated in (**f**) within the three-dimensional structure of M1.

Antibodies against 565 peptides significantly increased with age, primarily from herpesviruses HSV-1, HSV-2, and EBV, which can reactivate throughout life^35^. These associations were not due to cross-reactivity, as supported by AVARDA (Extended Data Fig. 2a), and were replicated in the EIP cohort for HSV-1 and EBV (Extended Data Fig. 2b-d). The strongest age effects were observed for antibodies targeting the US6 gene product of HSV-1, the surface protein glycoprotein D (Extended Data Fig. 2e), as well as various EBV proteins including EBNA-3, −4, and −6 (Extended Data Fig. 2f). Both peptide-level *Z*-scores and AVARDA breadth scores also showed positive associations with age for hepatitis A virus (HAV) and aichi virus A (Fig. 2a and Extended Data Fig. 2a), the latter being a kobuvirus initially isolated during a 1989 gastroenteritis outbreak in Japan that has subsequently been detected in Europe^36,37^. Conversely, antibodies against 766 peptides significantly decrease with age, primarily involving rhinoviruses, enteroviruses, and adenoviruses (Fig. 2a). After accounting for cross-reactivity with AVARDA, antibodies against rhinoviruses A and B, enterovirus B and C, and adenovirus D showed a significant decrease with age (Extended Data Fig. 2a), suggesting higher exposure in younger individuals and/or faster antibody waning in older adults.

Interestingly, antibodies against different IAV peptides strongly increase or decrease with age (Fig. 2a,b). Antibodies from younger individuals primarily target amino acid positions 1-100 and 300-400 of hemagglutinin (HA), which are part of the highly antigenic globular head of HA^38^, whereas older individuals preferentially target positions 450-550, which are part of the HA stalk domain (Fig. 2c-e). A similar pattern was observed for the IAV matrix protein 1 (MP1), with younger individuals more frequently targeting positions 200-250 and older individuals targeting positions 150-200 (Fig. 2f,g). These differences were not driven by age-related variations in exposure to different IAV subtypes, as both positive and negative associations were observed within the same IAV subtypes for HA and MP1 (Fig. 2c,f). Furthermore, although past flu vaccination was associated with higher total anti-IAV antibody titers in the MI cohort (*P* = 2.07 × 10^-14^), age was only weakly associated with vaccination (logistic regression *P* = 0.033), supporting the view that vaccination does not contribute to the observed patterns. Notably, the AVARDA breadth score for IAV was not associated with age (Extended Data Fig. 2a), as it aggregates peptides with opposite age effects (Methods). Together, these results indicate that epitope specificity of anti-IAV humoral responses varies with age.

The effects of sex on the antibody repertoire were moderate compared to those of age: 330 peptides showed significantly higher antibody levels in women and 236 in men (Extended Data Fig. 3a). While associated peptides originated from various viruses, AVARDA analysis supported higher reactivity in women for antibodies against CMV, HHV-6A, and HHV-6B (Extended Data Fig. 3b). These results suggest that women have higher exposure and/or stronger humoral responses to herpesviruses compared to men, in contrast to bacterial infections, which affect the antibody levels similarly in both sexes^4^. We observed that antibodies of women and men tend to target different IAV and IBV proteins, with women more often targeting the HA protein (Extended Data Fig. 3c,d). Given the similar flu vaccination rates between women and men in the MI cohort (20.2% *vs.* 18.6%, respectively; logistic regression *P* = 0.51), these findings suggest inherent sex differences in humoral responses against influenza viruses.

### Antibody profiles markedly differ according to population of origin

To investigate how geographical differences in pathogen exposure affect the antiviral antibody repertoire, we leveraged the EIP cohort, comprising individuals born in Central Africa (AFB) or Europe (EUB). While all samples were collected in Belgium, AFB had relocated to Europe shortly before sample collection (2.45 years before, on average^39^), implying that differences with EUB may reflect variations in early-life exposures and/or genetic ancestry. We observed marked population differences in antibody repertoires (Fig. 3a). Specifically, antibody levels against 898 viral peptides were increased in EUB, predominantly from rhinoviruses, adenoviruses, and IAV (*P*adj < 0.05), although significance was weak when considering the AVARDA scores (*P*adj > 0.001). In contrast, higher antibody reactivity in AFB was observed for 647 peptides, of which 61% were related to herpesviruses. The higher reactivity of AFB to herpesviruses was strongly supported by AVARDA for antibodies against CMV (*P*adj = 1.29 × 10^-19^), HHV-6A (*P*adj = 6.18 × 10^-17^), HHV-6B (*P*adj = 1.34× 10^-10^), and HHV-8 (*P*adj = 6.93 × 10^-20^) (Extended Data Fig. 4a), confirming previous studies^33,40,41^. Notably, anti-HHV-8 antibodies were significantly higher in AFB for 68 out of 70 peptides (Extended Data Fig. 4b). Similarly, reactivity against 108 out of 123 CMV peptides was greater in AFB, with the most significant antibodies targeting RL12, UL32/pp150, and UL139 (Extended Data Fig. 4c).

**Fig. 3:**
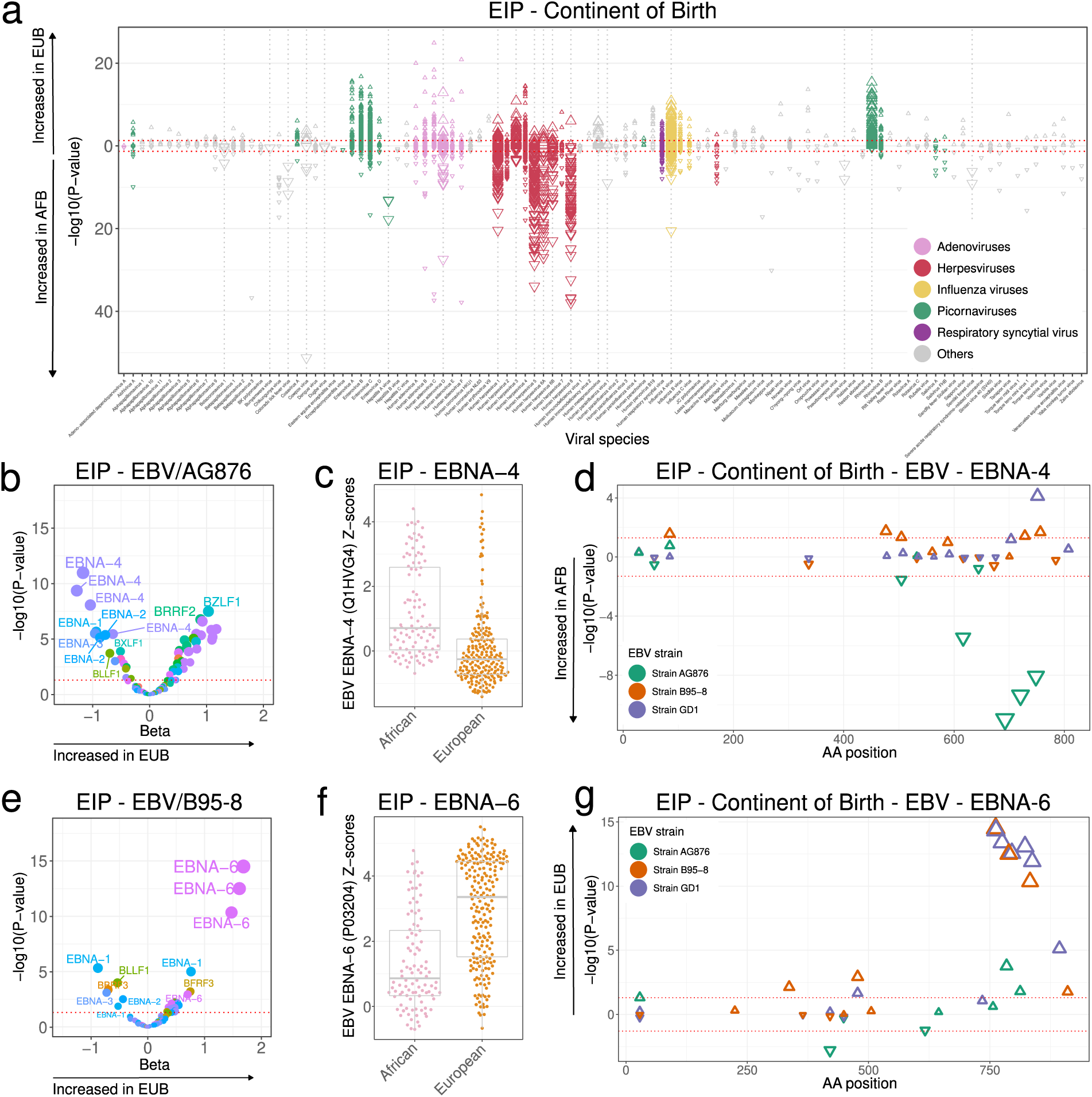
The antiviral antibody repertoire in relation to the continent of birth. **a**, – log10(adjusted *P*-values) and direction of associations between all public peptide Z-scores and continent of birth in the EIP cohort, separated by viral species. AFB and EUB indicate Belgian individuals born in Central Africa and Europe, respectively. The dashed gray vertical lines indicate viruses for which the AVARDA breadth score is significantly associated with continent of birth. **b**, –log10(adjusted *P*-values) against effect sizes of associations between continent of birth and peptide *Z*-scores from the EBV AG876 strain in the EIP cohort. Colors indicate the viral protein. **c**, Scatter plot of antibody reactivity against the most significant EBNA-4 peptide (Uniprot ID: Q1HVG4) from the EBV AG876 strain, categorized by continent of birth, **d**, Amino-acid positions of the midpoint of public EBNA-4 peptides associated with continent of birth within the full EBV EBNA-4 protein for the MI cohort. The significance and direction of associations with age are indicated on the y-axis and by the direction of triangles, respectively. The triangle colors indicate EBV strain. **e**, –log10(adjusted *P*-values) against effect sizes of associations between continent of birth and peptide *Z*-scores from the EBV B95−8 strain in the EIP cohort. Colors indicate the viral protein. **f**, Scatter plot of antibody reactivity against the most significant EBNA-6 peptide (Uniprot ID: P03204) from EBV B95−8, categorized by continent of birth, **g**, Amino-acid positions of EBV peptides in the EBNA-6 protein associated with continent of birth in the EIP cohort. Amino-acid positions of the midpoint of all public EBNA-6 peptides associated with continent of birth within the full EBV EBNA-6 protein for the MI cohort. The significance and direction of associations with age are indicated on the y-axis and by the direction of triangles, respectively. The triangle color indicates the EBV strain.

Antibody reactivity also differed between populations for epitopes from the same virus species. While overall reactivity to EBV was similar between AFB and EUB (*P*adj > 0.05; Extended Data Fig. 4a), the two groups targeted different EBV peptides (Fig. 3a). Antibodies from AFB more frequently targeted the viral protein EBNA-4, whereas those from EUB preferentially targeted EBNA-6 (Fig. 3b,e). The four EBNA-4 peptides most associated with African origin are located between amino acid positions 600-800 and derive from the AG876 strain, a type-2 EBV strain prevalent in Africa^42^ (Fig. 3b-d). Conversely, EBNA-6 peptides associated with European origin are found between amino acid positions 750-850 and derive from the GD1 and B95-8 cosmopolitan strains (Fig. 3e-g). These findings suggest that differences in epitope specificity between populations likely result from past exposure to different EBV strains. Similarly, antibodies against IAV from AFB primarily targeted NP from H1N1, whereas those from EUB favored HA from H3N2 (Extended Data Fig. 4d-f).

Collectively, these results reveal population disparities in antibody reactivity against epitopes of common viruses, highlighting the limitation of using single antigens to assess seroprevalence in global epidemiological studies.

### Smoking exerts strong yet reversible effects on antibody reactivity against rhinoviruses

To gain a more comprehensive understanding of the effects of non-genetic factors on the antiviral antibody repertoire, we leveraged the MI cohort to search for associations with a curated list of 108 variables assessing socio-economic status (SES), health-related habits, medical history, and disease-related biomarkers, while controlling for age and sex (Table S2, Methods). Besides weak associations with SES and a few health biomarkers (Fig. 4a; Table S3; Supplementary Note), the only strongly significant associations were found for tobacco smoking, which was associated with 134 peptides (Fig. 4a,b), primarily from rhinoviruses A and B and enteroviruses A-D. AVARDA analysis confirmed the significant association between cigarette consumption and antibodies targeting rhinoviruses A and B (*Padj* = 1.99 × 10^-4^). Rhinoviruses are prevalent causes of the common cold, which is more frequent and severe in smokers, although the underlying physiological mechanisms are debated^43,44^.

**Fig. 4:**
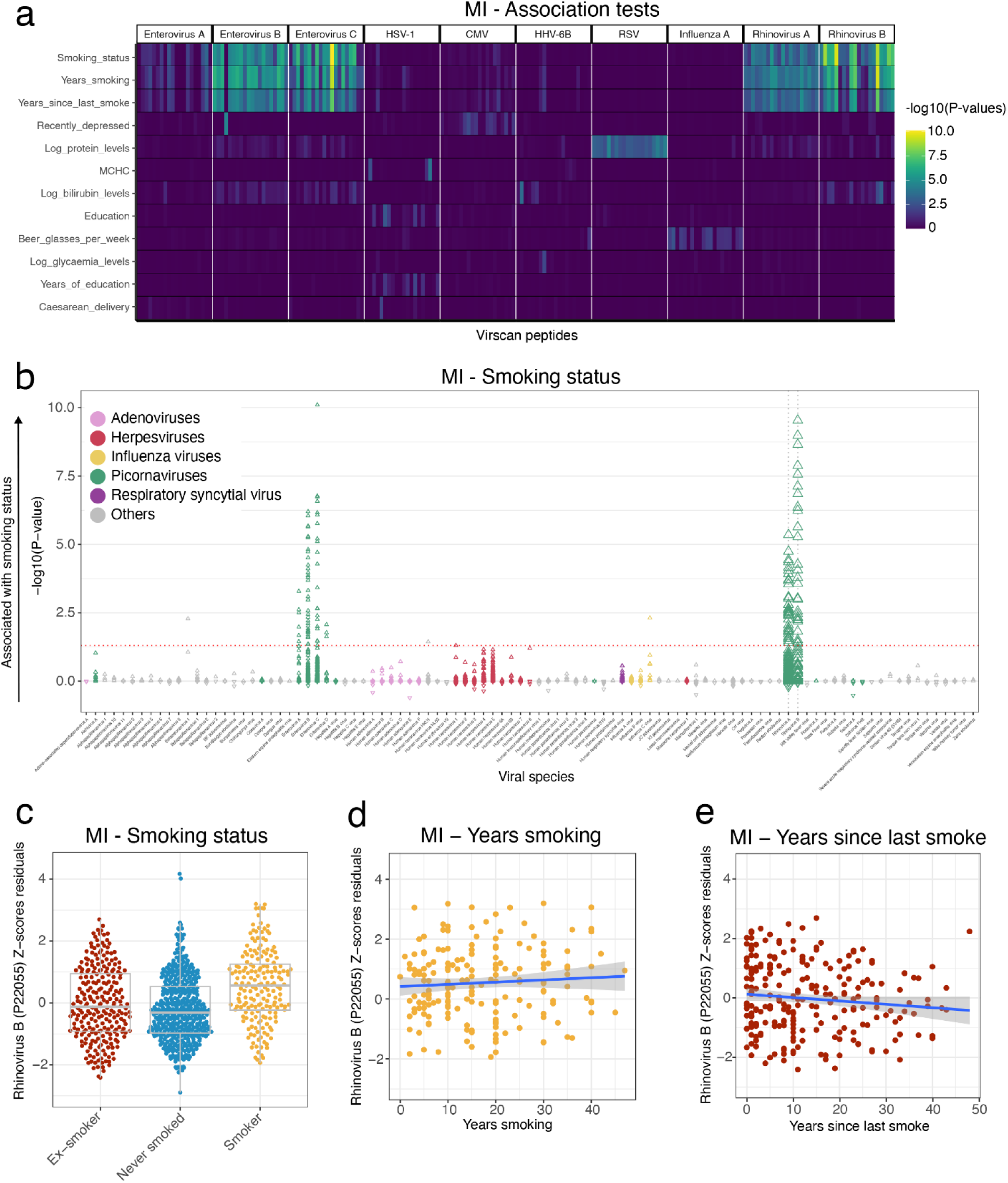
Tobacco smoking elicits strong, reversible effects on antiviral antibody responses. **a**, –log10(adjusted *P*-values) for associations between public peptide Z-scores and health- and lifestyle-related variables. Only the 20 most significant peptides from the ten viruses with the most significant associations are shown. Only variables with an association of *P*adj < 0.01 are shown. **b,** –log10(adjusted *P*-values) and direction of associations between all public peptide *Z*-scores and smoking status in the MI cohort, separated by viral species. The direction indicates positive or negative association with smoking compared to non-smokers. The dashed gray vertical lines indicate viruses for which the AVARDA breadth score is significantly associated with smoking status. **c**, Antibody reactivity for the rhinovirus B peptide most significantly associated with smoking status, categorized by smoking status. **d**, Antibody reactivity for the rhinovirus B peptide most significantly associated with smoking status, as a function of years of smoking in active smokers. **e**, Antibody reactivity for the rhinovirus B peptide most significantly associated with smoking status, as a function of years since last smoking in former smokers. **d**, **e**, The blue line indicates the linear regression line, and the shaded area the 95% confidence intervals.

The peptide most significantly associated with smoking originates from a rhinovirus B polyprotein containing capsid proteins, with antibody levels against it showing a large increase in smokers (*P*adj = 3.24 × 10^-10^; Fig. 4c). We found that anti-rhinovirus B reactivity was not associated with smoking duration in active smokers (*P* = 0.454) (Fig. 4d), suggesting constant, non-cumulative exposure to rhinoviruses. Interestingly, ex-smokers exhibited similar levels of reactivity compared to individuals who never smoked (*P* = 0.059; Fig. 4c). Accordingly, anti-rhinovirus B antibodies decreased with years after quitting smoking in former smokers (*P* = 5.97 × 10^-3^; Fig. 4e). These findings collectively indicate that smoking exerts a strong, yet reversible, effect on the antibody repertoire against rhinoviruses.

### Germline variants in immunoglobulin genes shape the antiviral antibody repertoire

To identify genetic factors affecting the antiviral antibody repertoire, we conducted a GWAS of *Z*-scores for the 2,608 public peptides in the MI cohort, by testing for associations with 5,699,237 imputed common SNPs^45^ while controlling for age, sex, and genetic structure (Methods). The EIP cohort served as a replication cohort. Given the incomplete coverage of B-cell receptor loci by the imputed SNPs, we performed next-generation sequencing of the *IGH*, *IGK*, and *IGL* genes in all MI donors at a ∼35× depth of coverage, generating an additional 30,503 common variants (Methods). We detected strong genome-wide significant associations for 225 viral peptides at four independent loci, including *HLA*, *FUT2*, *IGH*, and *IGK* genes (Fig. 5a; Tables 1 and S4).

**Fig. 5:**
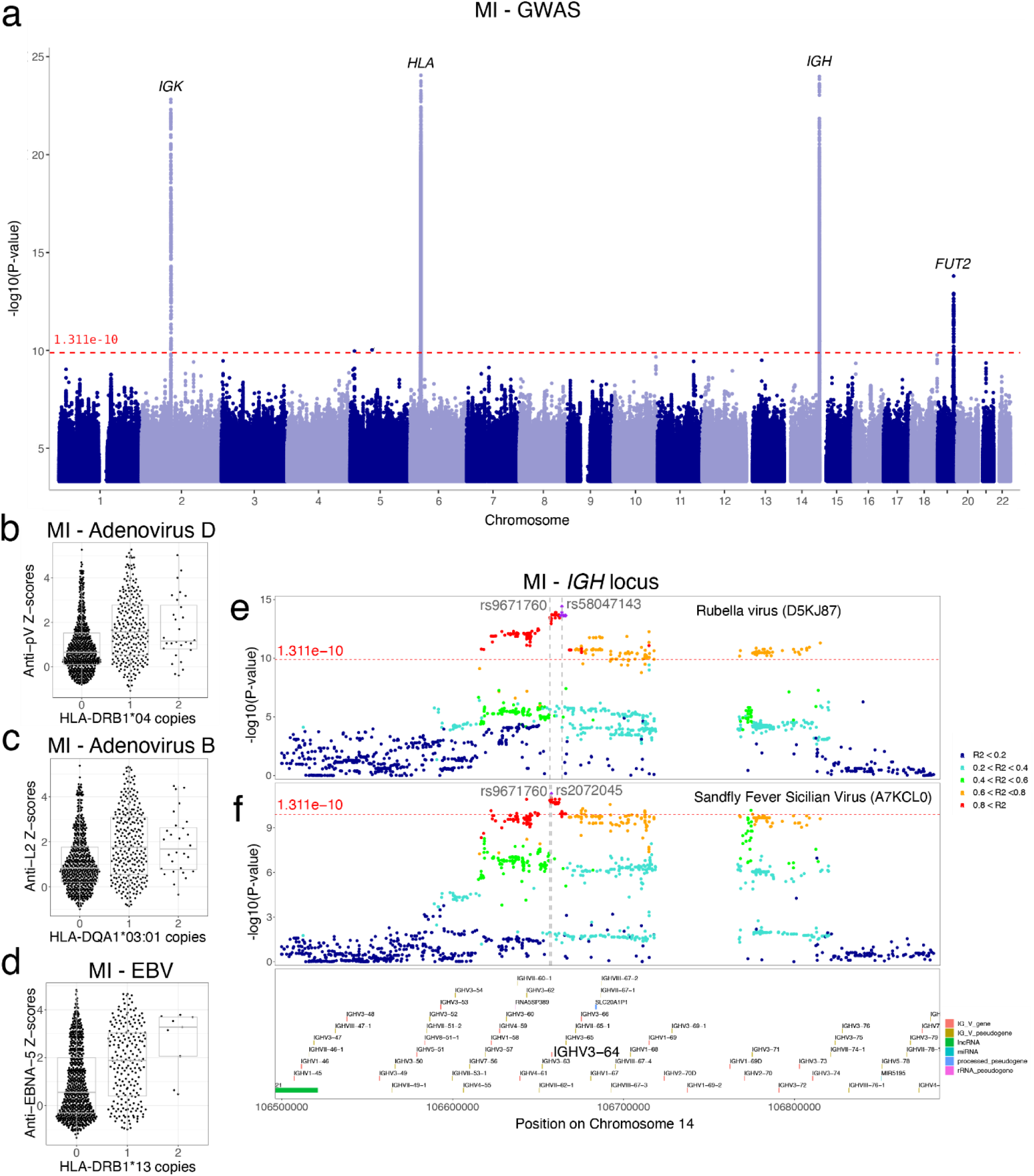
Genome-wide association study of antibody reactivity against public peptides. **a**, Manhattan plot of associations between all 2,608 public peptides and common human genetic variants (MAF > 5%) in the MI cohort. Only results with *P* < 0.005 are displayed. The red dashed line indicates the significance threshold (*P* < 1.31 × 10^-10^), determined by permutations. The top hit of each peak is annotated with the closest gene or gene locus. **b**, Antibody reactivity against the pV protein of adenovirus D, as a function of the number of copies of the *HLA-DRB1*\*04 allele. **c**, Antibody reactivity against the L2 protein of adenovirus B, as a function of the number of copies of the *HLA-DQA1*\*03:01 allele. **d**, Antibody reactivity against the EBNA-5 protein of EBV, as a function of the number of copies of the *HLA-DRB1*\*13 allele. **e,f**, LocusZoom plots for the associations between *IGH* variants and antibody reactivity against (**e**) the rubella virus (UniProt ID: D5KJ87) and (**f**) the sandfly fever Sicilian virus (UniProt ID: A7KCL0). The variant most significantly associated with antibody reactivity and the closest guQTL variant (rs9671760) are indicated by gray vertical lines. *IGHV* segment locations are indicated at the bottom, and the V-segment targeted by the guQTL variant (*IGHV3-64*) is highlighted.

We found significant associations between *HLA* variants and antibody reactivity against 112 peptides from 15 viruses, including EBV, HSV-1, and adenoviruses A-F, consistent with prior studies^2,4,10,11,14–16^ and replicated in the EIP cohort (*P*rep < 0.05; Tables 1 and S4). To account for linkage disequilibrium (LD) among *HLA* variants and enable comparisons with previous disease studies, we imputed *HLA* alleles from genotype data and tested for associations between peptide *Z*-scores and allele dosages (Methods). This analysis revealed 85 associations (Table S5), including *HLA-DRB1*\*04 and *HLA-DQA1*\*03:01 with adenovirus peptides (*P* < 2.3 × 10^-15^; Fig. 5b,c) and *HLA-DRB1*\*13 with EBV peptides (*P* = 7.5 × 10^-19^; Fig. 5d). Notably, these alleles have previously been associated with increased risk for type 1 diabetes and rheumatoid arthritis^46^, providing a potential explanation for the link between these immune diseases and EBV and adenovirus infections^47–49^.

Variants near *FUT2* were associated with antibodies against norovirus peptides (*P* = 1.10 × 10^-10^; Extended Data Fig. 5a). Mutations in *FUT2* determine the non-secretor phenotype, which is known to confer resistance to norovirus infection^50^ and susceptibility to type 1 diabetes and inflammatory bowel disease^51,52^. The most significant variants include rs601338 (*P* = 2.01 × 10^-10^), the *FUT2* stop mutation that commonly determines the non-secretor status^53^, the protective allele being associated with lower anti-norovirus antibody levels. Variants in strong LD with rs601338 also showed significant associations in the EIP cohort (*P*rep = 5.98 × 10^-9^; *r²* = 0.998). Additionally, we identified a novel association between variants in near-complete LD (*r²* = 0.995) with rs601338 and antibodies against two salivirus strains in both the MI (*P* < 1.58 × 10^-14^) and EIP (*P*rep < 1.36 × 10^-10^) cohorts (Extended Data Fig. 5b). Saliviruses, first discovered in 2009 in diarrheal samples, are known to cause gastroenteritis^54^, although their target cells and entry mechanisms remain unknown. The associations between the *FUT2* non-secretor status and anti-salivirus antibodies are unlikely to result from cross-reactivity with norovirus peptides, as their respective *Z*-scores were not correlated (Extended Data Fig. 5c,d).

Genetic variation within the *IGH* locus was associated with 107 peptides from 21 viruses (Fig. 5a; Tables 1 and S4). This genomic region encodes the heavy chain of the antibody molecule and has previously been associated with antibody levels against various bacteria, as well as IAV and norovirus^4^. Our analyses expanded these findings, by identifying new associations with herpesviruses (HSV-2, EBV, CMV, HHV-6), RSV, IAV, HBV, coronavirus NL63, rubella virus, sandfly fever Sicilian virus, enteroviruses, and rhinoviruses. Interestingly, several newly identified GWAS variants influence *IGHV* clonal gene usage by V(D)J somatic recombination, assessed by AIRR-sequencing in a previous study^55^. For example, we found that a variant associated with antibodies against the rubella virus (rs1024350, *P* = 1.90 × 10^-11^) and suggestively associated with IAV (*P* = 5.38 × 10^-10^) affects *IGHV1-69* usage^55^ (*P* = 1.14 × 10^-16^). *IGHV1-69* usage is known to partially determine the quality of anti-influenza antibodies^56^. Another variant, rs9671760, which we found associated with antibodies against the rubella virus (*P* = 3.34 × 10^-14^; Fig 5e) and the sandfly fever Sicilian virus (*P* = 1.46 × 10^-11^; Fig 5f), regulates *IGHV3-64* usage^55^ (*P* = 1.32 × 10^-8^).

The fourth genome-wide significant locus revealed a novel association between *IGK*, which encodes the κ light chain of antibodies, and antibody levels targeting adenovirus B peptides (*P* = 1.51 × 10^-23^) (Extended Data Fig. 5e). Together, these findings underscore the broad impact of host genetic factors, including germline mutations in immunoglobulin genes, on humoral immune responses to multiple viruses.

### Demographic and genetic factors differentially affect reactivity across viral epitopes

Finally, to assess the relative contributions of demographic (non-genetic) and genetic factors to the antibody repertoire, we estimated the proportion of variance explained by age, sex, smoking, and GWAS lead variants for the 2,608 public peptides. Together, these factors explained an average of 7.39% (range: [0.91% – 25.50%]) of inter-individual variation in antibody reactivity (Fig 6a). Demographic factors explained 3.81% (range: [0.007% – 20.68%]) of the variance, while genetic factors contributed to 3.44% (range: [0.48% – 23.02%]). These proportions varied substantially across viruses, consistent with earlier findings (Extended Data Fig. 6a,b). For example, antibody levels against rhinovirus peptides were predominantly affected by age (Fig. 2a), those against CMV by sex (Extended Data Fig. 3a), and those against EBV by genetic variation (Table 1).

**Fig. 6:**
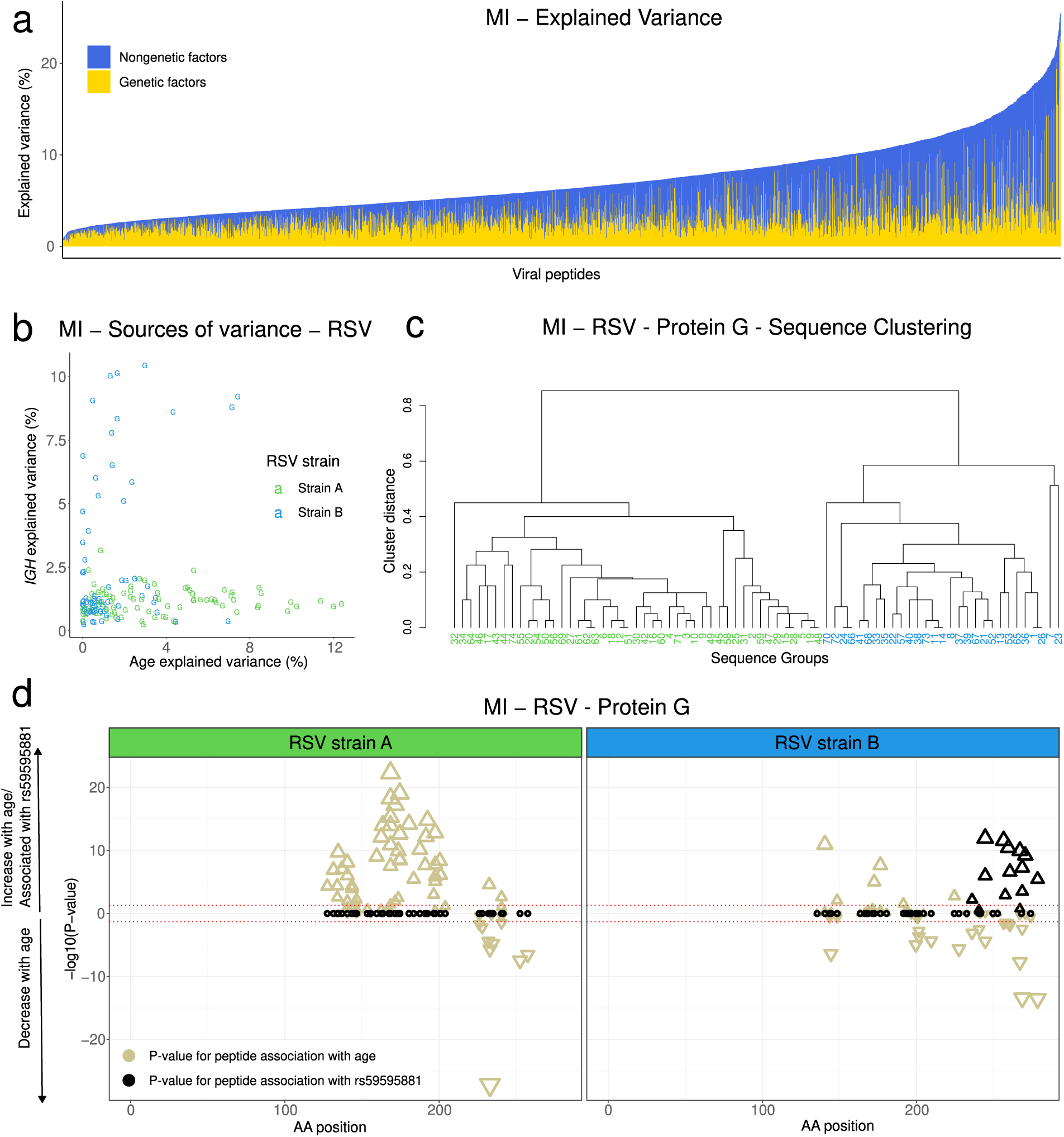
Variance in antiviral antibody reactivity explained by demographic and genetic factors. **a**, Proportion of variance explained by demographic (i.e., age, sex, and smoking) and genetic factors for antibody reactivity against 2,608 public peptides in the MI cohort. Peptides are sorted by total variance explained. **b**, Variance explained by age and *IGH* genetic variation for RSV protein G peptides in the MI cohort, colored according to RSV strain as in (**c)**. **c**, Hierarchical clustering of peptide sequences from RSV protein G, separating peptides affiliated to the RSV A (green) and B (blue) strains. **d**, Amino-acid positions of the midpoint of protein G peptides associated with continent of birth within the full RSV protein G for the MI cohort. *P*-values for the association with age (beige) and the most significant *IGH* variant (black) are indicated, separated by RSV strain. The significance and direction of associations are indicated on the y-axis and by the direction of triangles, respectively.

**Table 1.**
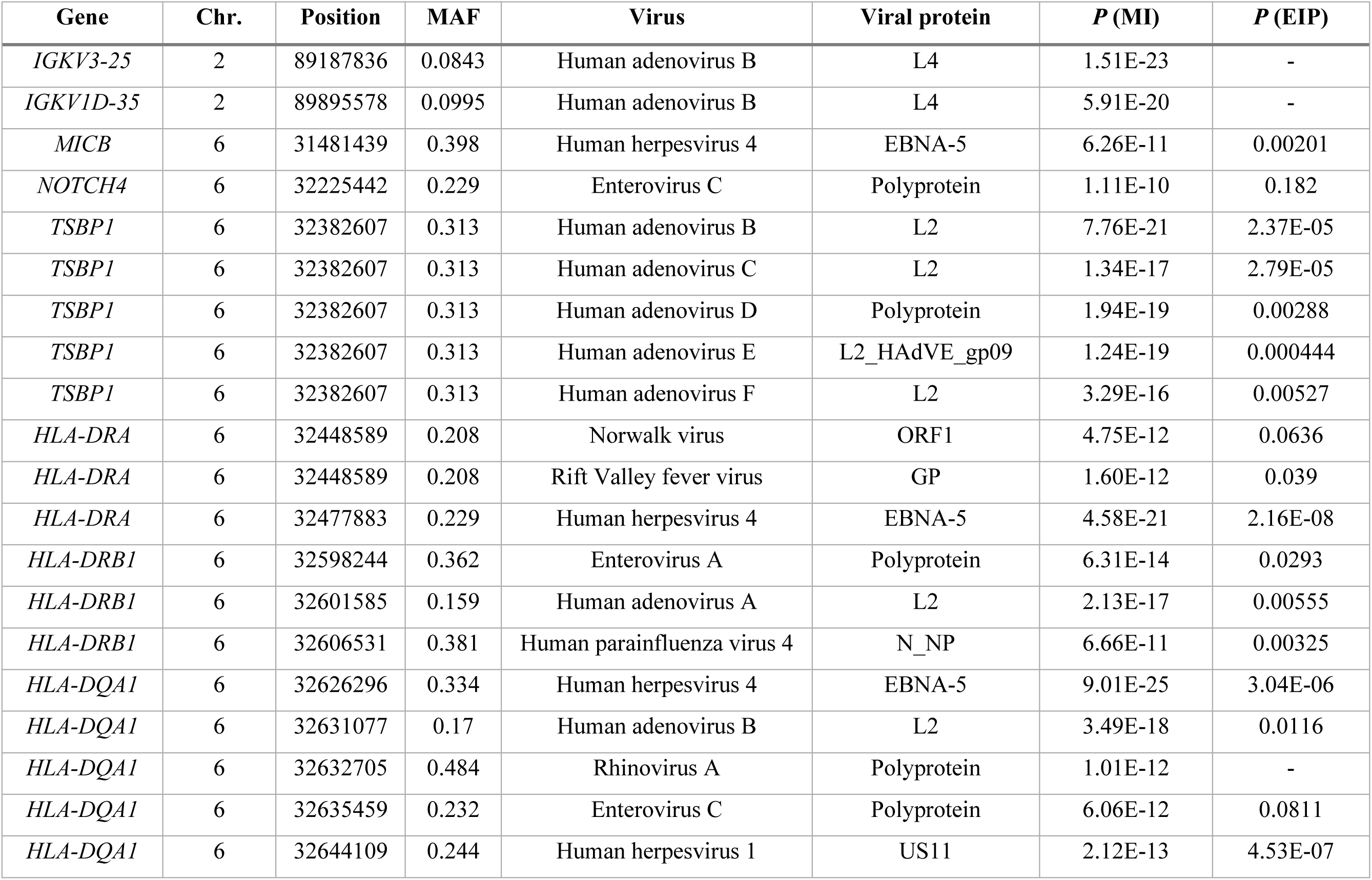
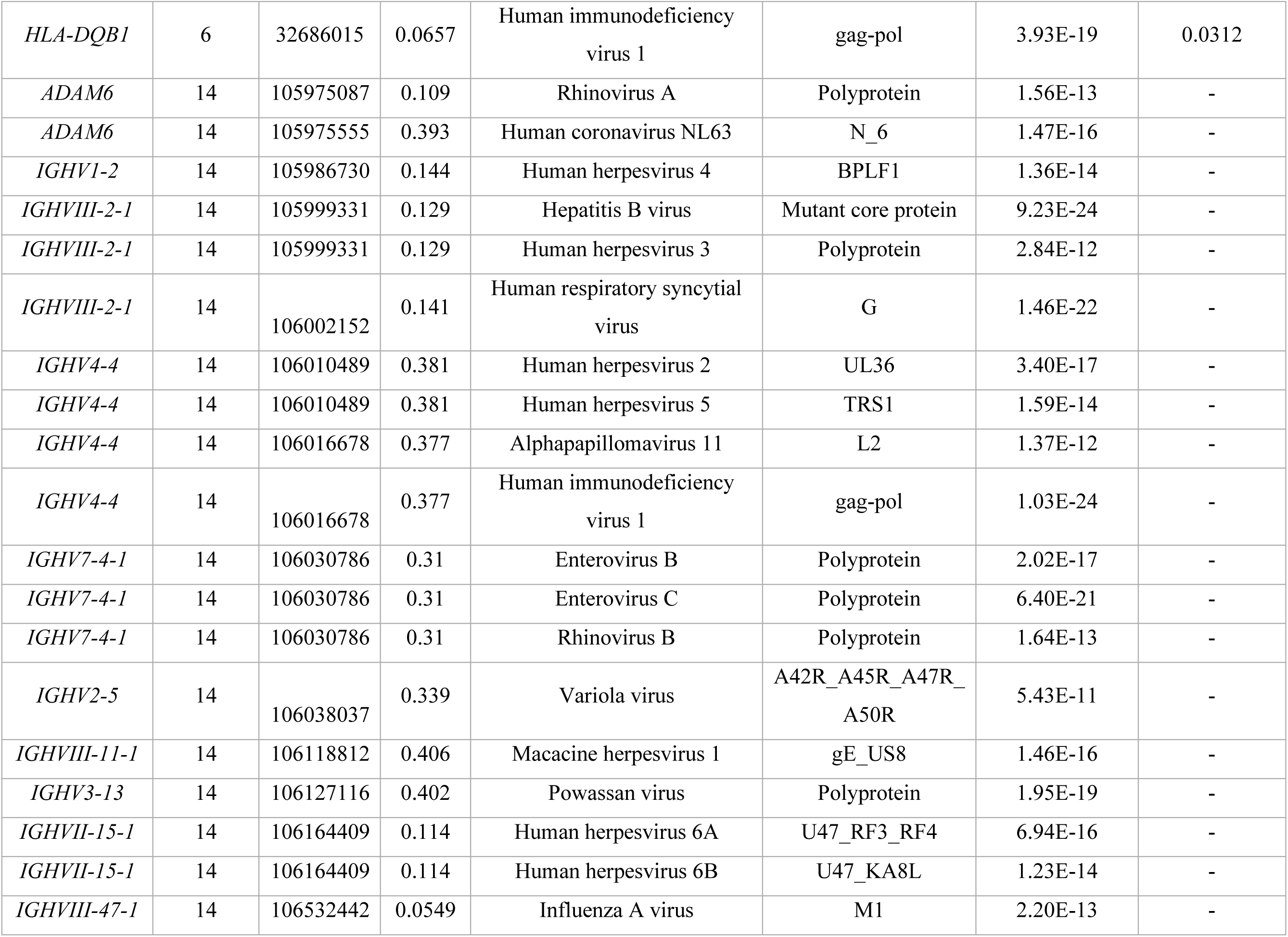
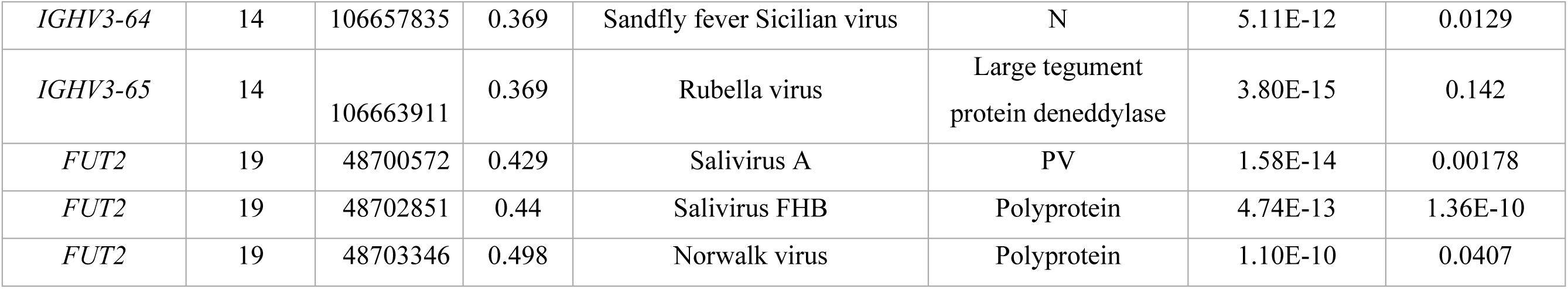
Genome-wide significant associations between human genetic variants and the antibody repertoire. Only the most significant variant within a 1-Mb window centered on genome-wide significance hits is shown for each associated virus.

| Gene | Chr. | Position | MAF | Virus | Viral protein | *P* (MI) | *P* (EIP) |
| --- | --- | --- | --- | --- | --- | --- | --- |
| *IGKV3-25* | 2 | 89187836 | 0.0843 | Human adenovirus B | L4 | 1.51E-23 | - |
| *IGKV1D-35* | 2 | 89895578 | 0.0995 | Human adenovirus B | L4 | 5.91E-20 | - |
| *MICB* | 6 | 31481439 | 0.398 | Human herpesvirus 4 | EBNA-5 | 6.26E-11 | 0.00201 |
| *NOTCH4* | 6 | 32225442 | 0.229 | Enterovirus C | Polyprotein | 1.11E-10 | 0.182 |
| *TSBP1* | 6 | 32382607 | 0.313 | Human adenovirus B | L2 | 7.76E-21 | 2.37E-05 |
| *TSBP1* | 6 | 32382607 | 0.313 | Human adenovirus C | L2 | 1.34E-17 | 2.79E-05 |
| *TSBP1* | 6 | 32382607 | 0.313 | Human adenovirus D | Polyprotein | 1.94E-19 | 0.00288 |
| *TSBP1* | 6 | 32382607 | 0.313 | Human adenovirus E | L2_HAdVE_gp09 | 1.24E-19 | 0.000444 |
| *TSBP1* | 6 | 32382607 | 0.313 | Human adenovirus F | L2 | 3.29E-16 | 0.00527 |
| *HLA-DRA* | 6 | 32448589 | 0.208 | Norwalk virus | ORF1 | 4.75E-12 | 0.0636 |
| *HLA-DRA* | 6 | 32448589 | 0.208 | Rift Valley fever virus | GP | 1.60E-12 | 0.039 |
| *HLA-DRA* | 6 | 32477883 | 0.229 | Human herpesvirus 4 | EBNA-5 | 4.58E-21 | 2.16E-08 |
| *HLA-DRB1* | 6 | 32598244 | 0.362 | Enterovirus A | Polyprotein | 6.31E-14 | 0.0293 |
| *HLA-DRB1* | 6 | 32601585 | 0.159 | Human adenovirus A | L2 | 2.13E-17 | 0.00555 |
| *HLA-DRB1* | 6 | 32606531 | 0.381 | Human parainfluenza virus 4 | N_NP | 6.66E-11 | 0.00325 |
| *HLA-DQA1* | 6 | 32626296 | 0.334 | Human herpesvirus 4 | EBNA-5 | 9.01E-25 | 3.04E-06 |
| *HLA-DQA1* | 6 | 32631077 | 0.17 | Human adenovirus B | L2 | 3.49E-18 | 0.0116 |
| *HLA-DQA1* | 6 | 32632705 | 0.484 | Rhinovirus A | Polyprotein | 1.01E-12 | - |
| *HLA-DQA1* | 6 | 32635459 | 0.232 | Enterovirus C | Polyprotein | 6.06E-12 | 0.0811 |
| *HLA-DQA1* | 6 | 32644109 | 0.244 | Human herpesvirus 1 | US11 | 2.12E-13 | 4.53E-07 |
| *HLA-DQB1* | 6 | 32686015 | 0.0657 | Human immunodeficiency virus 1 | gag-pol | 3.93E-19 | 0.0312 |
| *ADAM6* | 14 | 105975087 | 0.109 | Rhinovirus A | Polyprotein | 1.56E-13 | - |
| *ADAM6* | 14 | 105975555 | 0.393 | Human coronavirus NL63 | N_6 | 1.47E-16 | - |
| *IGHV1-2* | 14 | 105986730 | 0.144 | Human herpesvirus 4 | BPLF1 | 1.36E-14 | - |
| *IGHVIII-2-1* | 14 | 105999331 | 0.129 | Hepatitis B virus | Mutant core protein | 9.23E-24 | - |
| *IGHVIII-2-1* | 14 | 105999331 | 0.129 | Human herpesvirus 3 | Polyprotein | 2.84E-12 | - |
| *IGHVIII-2-1* | 14 | 106002152 | 0.141 | Human respiratory syncytial virus | G | 1.46E-22 | - |
| *IGHV4-4* | 14 | 106010489 | 0.381 | Human herpesvirus 2 | UL36 | 3.40E-17 | - |
| *IGHV4-4* | 14 | 106010489 | 0.381 | Human herpesvirus 5 | TRS1 | 1.59E-14 | - |
| *IGHV4-4* | 14 | 106016678 | 0.377 | Alphapapillomavirus 11 | L2 | 1.37E-12 | - |
| *IGHV4-4* | 14 | 106016678 | 0.377 | Human immunodeficiency virus 1 | gag-pol | 1.03E-24 | - |
| *IGHV7-4-1* | 14 | 106030786 | 0.31 | Enterovirus B | Polyprotein | 2.02E-17 | - |
| *IGHV7-4-1* | 14 | 106030786 | 0.31 | Enterovirus C | Polyprotein | 6.40E-21 | - |
| *IGHV7-4-1* | 14 | 106030786 | 0.31 | Rhinovirus B | Polyprotein | 1.64E-13 | - |
| *IGHV2-5* | 14 | 106038037 | 0.339 | Variola virus | A42R_A45R_A47R_A50R | 5.43E-11 | - |
| *IGHVIII-11-1* | 14 | 106118812 | 0.406 | Macacine herpesvirus 1 | gE_US8 | 1.46E-16 | - |
| *IGHV3-13* | 14 | 106127116 | 0.402 | Powassan virus | Polyprotein | 1.95E-19 | - |
| *IGHVII-15-1* | 14 | 106164409 | 0.114 | Human herpesvirus 6A | U47_RF3_RF4 | 6.94E-16 | - |
| *IGHVII-15-1* | 14 | 106164409 | 0.114 | Human herpesvirus 6B | U47_KA8L | 1.23E-14 | - |
| *IGHVIII-47-1* | 14 | 106532442 | 0.0549 | Influenza A virus | M1 | 2.20E-13 | - |
| *IGHV3-64* | 14 | 106657835 | 0.369 | Sandfly fever Sicilian virus | N | 5.11E-12 | 0.0129 |
| *IGHV3-65* | 14 | 106663911 | 0.369 | Rubella virus | Large tegument<br>protein deneddylase | 3.80E-15 | 0.142 |
| *FUT2* | 19 | 48700572 | 0.429 | Salivirus A | PV | 1.58E-14 | 0.00178 |
| *FUT2* | 19 | 48702851 | 0.44 | Salivirus FHB | Polyprotein | 4.74E-13 | 1.36E-10 |
| *FUT2* | 19 | 48703346 | 0.498 | Norwalk virus | Polyprotein | 1.10E-10 | 0.0407 |

We also observed substantial variation in the factors explaining the variance of peptide *Z*-scores within the same virus. For example, the variance of antibody reactivity to the HA protein of IAV was predominantly explained by age, whereas anti-M1 antibodies were primarily affected by *IGH* genetic variation (Extended Data Fig. 6c). Similarly, anti-EBV antibodies targeting the EBNA-5 protein were strongly influenced by *HLA* genotypes, while those targeting EBNA-4 and tegument proteins varied primarily because of age (Extended Data Fig. 6d).

Interestingly, a similar pattern was observed for anti-RSV antibodies, but at the level of a single protein: antibodies against different peptides of the immunogenic glycoprotein G^57^ were associated with either age or *IGH* genetic variants (Fig 6b). Age-associated peptides originate from RSV strain A, while *IGH*-associated peptides originate from strain B — two phylogenetic RSV lineages that differ substantially in the protein G sequence^58^ (Fig. 6c). Specifically, age-associated antibodies primarily targeted amino acid positions 150-200 of protein G in RSV-A (Fig 6d), a pattern confirmed in the EIP cohort (Extended Data Fig. 6e) and a previous study^59^. In contrast, *IGH*-associated antibodies were predominantly directed at positions 225-275 of RSV-B (Fig 6d), indicating strain- and position-specific genetic effects. Overall, these findings indicate that the effects of non-genetic and genetic factors largely differ among viruses, viral strains, proteins, and epitopes targeted by the human antibody repertoire.

## Discussion

In this study, we generated a comprehensive dataset of blood plasma antibody levels against more than 97,000 viral peptides, providing a valuable resource to investigate the factors — intrinsic, environmental, and genetic — that affect the antibody repertoire in healthy adults. All results can be explored via a dedicated web-based browser (http://mirepertoire.pasteur.cloud/). Among these factors, age had the most profound and widespread effect on antibody reactivity. Age-related increases in antibody response may reflect cumulative exposure in older adults (e.g., HAV and aichi virus A), reactivation of latent viruses (e.g., HSV-1, HSV-2, EBV, and CMV), or reinfections by viruses causing recurrent infections (e.g., IAV, IBV, and RSV). Conversely, age-related decreases may reflect higher exposure during young adulthood and rapid antibody waning (e.g., rhinoviruses A-C and enteroviruses B and C).

Importantly, our study reveals that aging is associated with differential epitope recognition for the same viral protein. Anti-IAV antibodies of younger and older adults target different domains of the same IAV proteins, a phenomenon observed across IAV subtypes and for the IAV M1 protein, which is not typically targeted by flu vaccines. This suggests that the observed differences are not solely attributable to age-related disparities in natural or vaccine-induced exposure to diverse viral strains. Alternatively, certain structural domains of viral proteins may be less accessible to antibodies, necessitating multiple reinfections to elicit antibodies against them. This hypothesis has been proposed to explain age-related differences in neutralizing antibody titers against the globular head and stalk domains of the IAV HA protein^60,61^. We propose that age-dependent antigenic specificity, observed here for the first time across several IAV proteins, may be more widespread than previously recognized. Similarly, we show that sex influences epitope specificity, with women’s antibodies preferentially targeting the HA protein of IAV and IBV, relative to men, while men’s antibodies disproportionally target NP and M1. Further studies are needed to elucidate the underlying mechanisms and their implications for age- and sex-related differences in the risk of influenza infection and vaccine response.

Antibody profiles also vary markedly according to the continent of birth, likely due to differences in viral exposure^27^. We observed that antibodies from individuals born in Central Africa or Europe target different EBV proteins, suggesting that regional variations in EBV strains^33,34^ contribute to population differences in antibody responses at the epitope level. Among the environmental factors affecting the antibody repertoire, we identified a strong association between smoking and anti-rhinovirus antibodies, consistent with the higher risk of smokers for the common cold compared to non-smokers. Notably, we observed similar antibody levels against rhinoviruses in ex-smokers and never-smokers, indicating that altered viral clearance and/or heightened exposure in smokers is reversible upon smoking cessation.

Finally, our GWAS confirms that *HLA* and *IGH* affect antibody levels against a range of viruses^2,4,10,11,14–16^, and largely expands the list of associated viruses, by revealing novel associations with herpesviruses 2-6, RSV, HBV, rhinoviruses, enteroviruses, coronavirus N63, and rubella virus. Sequencing of the immunoglobulin genes was critical in discovering associations with the *IGH* locus, as well as the new association with *IGK*, since SNP arrays do not cover these complex regions. We also identified a strong association between antibodies against the recently discovered and poorly understood saliviruses and *FUT2*, a gene previously linked to norovirus infection, suggesting that saliviruses may utilize similar infection mechanisms as noroviruses.

Several genetic variants identified in our study as associated with increased humoral responses against viruses have previously been linked to higher risk of autoimmune diseases^46,51,52^. Patients with these diseases often show higher seroprevalence for common viruses, leading previous studies to suggest a causal role of these viral infections in autoimmunity^47–49^. However, our results suggest that associations between autoimmune conditions and antibody levels against viruses may instead result from a shared genetic etiology that affects both traits independently. Furthermore, our study supports the hypothesis of antagonistic pleiotropy, which posits that variants that once conferred resistance to infection now increase the risk for non-infectious immune diseases^63^. Consistent with this, the *HLA* and *FUT2* alleles associated with antiviral humoral responses have increased in frequency under natural selection in Europe over the past millennia^64^. Detailed sequencing-based studies in large biobanks are now required to determine the role of genetic variation in shaping the antibody repertoire in immune disorders.

This study has several limitations. First, while the VirScan library offers broad coverage, it is limited to linear peptides, potentially overlooking antibodies that bind to conformational epitopes. Additionally, antibody cross-reactivity between peptides introduces uncertainty in attributing results to specific viruses. We mitigated this risk by using AVARDA, although this method may also lead to false negatives. The extensive number of tests required to evaluate the entire peptide library, combined with the cohort size, may further increase false negatives.

Lastly, the PhIP-seq approach does not differentiate between neutralizing and non-neutralizing antibodies, which would require large-scale experimental studies. Despite these challenges, our study provides high-resolution insights into the widespread effects of age, sex, continent of birth, smoking, and genetics on the antibody repertoire. Crucially, it also uncovers how these factors differentially affect antibodies targeting specific epitopes within the same virus or viral protein, deepening our understanding of antibody generation and maintenance processes. We anticipate that our dataset and findings will prompt novel mechanistic studies of antiviral immunity, with the potential to advance vaccine and therapeutic strategies.

## Data Availability

The VirScan3 PhIP-seq raw and processed data generated in this study have been deposited in the Institut Pasteur data repository, OWEY, which can be accessed via the following link: https://dataset.owey.io/doi/10.48802/owey.84rn-jg72?version=1.1. All association statistics obtained in this study can be explored and downloaded from the web browser http://mirepertoire.pasteur.cloud/. The SNP array data can be accessed in the European Genome-Phenome Archive (EGA) with the accession code EGAS00001002460. All Milieu Intérieur datasets can be accessed by submitting a data access request to, the Milieu Intérieur data access committee (DAC). The DAC informs all the research participants of the data access request and grants data access if the request is consistent with the informed consent signed by the participants. In particular, research on Milieu Intérieur datasets is restricted to research on the genetic and environmental determinants of human variation in immune responses. Data access is typically granted two months after request submission.

## Acknowledgments

We acknowledge the help of the HPC Core Facility of Institut Pasteur for this work. A.O. was supported by a grant from the Wenner-Gren Foundation. This work received funding from the French government’s program ‘Investissement d’Avenir,’ managed by the *Agence Nationale de la Recherche* (reference 10-LABX-69-01).

## Author contributions

A.O., L.Q.-M., and E.P. conceived and developed the study. F.D. and B.C. prepared DNA samples. Z.T., C.P., and P.B. acquired VirScan data. D.D. and P.B. advised on experiments. E.B. and M.W. generated the Luminex-based serology data. E.C., I.L.A., and A.T. generated the Kappa-deleting recombination excision circles (KREC) data. A.O. performed all analyses, with contributions from A.J., M.R., D.L. and E.P. A.J. developed predictive algorithms. E.P. supervised analyses. A.O. and E.P. wrote the manuscript, with input from L.Q.-M. All authors discussed the results and contributed to the final manuscript.

## Declaration of interests

The authors declare no conflict of interest.

The Milieu Intérieur Consortium¶ is composed of the following team leaders: Laurent Abel (Hôpital Necker), Andres Alcover, Hugues Aschard, Philippe Bousso, Nollaig Bourke (Trinity College Dublin), Petter Brodin (Karolinska Institutet), Pierre Bruhns, Nadine Cerf-Bensussan (INSERM UMR 1163 – Institut Imagine), Ana Cumano, Christophe D’Enfert, Ludovic Deriano, Marie-Agnès Dillies, James Di Santo, Gérard Eberl, Jost Enninga, Jacques Fellay (EPFL, Lausanne), Ivo Gomperts-Boneca, Milena Hasan, Gunilla Karlsson Hedestam (Karolinska Institutet), Serge Hercberg (Université Paris 13), Molly A Ingersoll (Institut Cochin and Institut Pasteur), Olivier Lantz (Institut Curie), Rose Anne Kenny (Trinity College Dublin), Mickaël Ménager (INSERM UMR 1163 – Institut Imagine), Frédérique Michel, Hugo Mouquet, Cliona O’Farrelly (Trinity College Dublin), Etienne Patin, Antonio Rausell (INSERM UMR 1163 – Institut Imagine), Frédéric Rieux-Laucat (INSERM UMR 1163 – Institut Imagine), Lars Rogge, Magnus Fontes (Institut Roche), Anavaj Sakuntabhai, Olivier Schwartz, Benno Schwikowski, Spencer Shorte, Frédéric Tangy, Antoine Toubert (Hôpital Saint-Louis), Mathilde Touvier (Université Paris 13), Marie-Noëlle Ungeheuer, Christophe Zimmer, Matthew L. Albert (Octant Biosciences), Darragh Duffy§, Lluis Quintana-Murci§,

¶ unless otherwise indicated, partners are located at Institut Pasteur, Paris

§ co-coordinators of the Milieu Intérieur Consortium Additional information can be found at: https://www.milieuinterieur.fr/en/

---

**Supplementary Table 1.** Performance statistics for models predicting serostatus using VirScan peptide *Z*-scores

**Supplementary Table 2.** Demographic and lifestyle variables examined

**Supplementary Table 3.** *P*-values for the associations between demographic variables and 2,608 public peptide *Z*-scores

**Supplementary Table 4.** Association statistics between all genome-wide significant GWAS hits and 2,608 public peptide *Z*-scores

**Supplementary Table 5.** Association statistics for all significant *HLA* alleles

## Online Methods

### The Milieu Intérieur cohort

The Milieu Intérieur (MI) cohort comprises 1,000 healthy adults recruited to investigate genetic and non-genetic determinants of immune response variation^32^. Recruitment was conducted in Rennes (France) in 2012-2013, and individuals were selected based on a large set of relatively strict inclusion and exclusion criteria described elsewhere^32^. Of the 900 individuals reported in the present study, 453 are female, and 447 are male, ranging from 20 to 69 years of age. The study has been approved by the *Comité de Protection des Personnes — Ouest* 6 (Committee for the Protection of Persons) and by the French *Agence Nationale de Sécurité du Médicament* (ANSM). The study protocol, including inclusion and exclusion criteria for the Milieu Intérieur study, has been registered on ClinicalTrials.gov under the study ID NCT01699893.

### The EvoImmunoPop cohort

The EvoImmunoPop (EIP) cohort comprises 390 healthy adults recruited to investigate human population differences in immune responses. Recruitment was conducted in Ghent (Belgium) in 2012-2013. Of the 312 individuals reported in the present study, 100 individuals reported to be of Central African descent (AFB, age range 20 to 50 years), and 212 reported to be of European descent (EUB, age range 20 to 50 years). All EUB were born in Europe, whereas >90% of AFB were born in Cameroon or the Democratic Republic of Congo. AFB and EUB present no evidence of recent genetic admixture with populations originating from another continent, besides two AFB donors who present 22% of Near Eastern and 25% of European ancestries, respectively^33^. All individuals were negative for serological tests against human immunodeficiency virus, hepatitis B, or hepatitis C. The study has been approved by the Ethics Committee of Ghent University, the Ethics Board of Institut Pasteur (EVOIMMUNOPOP-281297), and the French authorities CPP, CCITRS, and CNIL.

### VirScan experimental protocol

To investigate the virus-specific and viral peptide-specific antibody profiles in the plasma of MI and EIP samples, we employed PhIP-Seq using the VirScan V3 library, a pathogen-epitope scanning method based on bacteriophage display and immuno-precipitation. The detailed protocol and VirScan library are described elsewhere^27,29,65^. Briefly, a library of linear peptides of 56 amino acids each was constructed to cover all UniProt protein sequences of viruses known to infect humans. Peptides were staggered along each protein sequence with an overlap of 28 amino acids. The phage library was inactivated and incubated with plasma samples normalized to total IgG concentration and controls (bead samples) to form IgG-phage immunocomplexes.

The immunocomplexes were then captured by magnetic beads, lysed, and sent to next-generation sequencing. Two replicates were performed for each individual to assess reproducibility.

### VirScan data preprocessing

Sequencing reads were processed as in ref.^28^, with some modifications. We utilized the bowtie2-samtools pipeline^66,67^ to map the sequencing reads of each sample to the bacteriophage library and count the number of reads for each viral peptide. Subsequently, the positivity of each peptide in plasma samples was determined by a binning strategy where read counts from blank controls were first used to group the peptides into hundreds of bins so that the counts form a uniform distribution within each bin. Then, the peptides from plasma samples were allocated into the pre-defined bins. *Z*-scores were calculated for each peptide from each plasma sample. The means and standard deviations used for the *Z*-score calculations were the same for each bin and were computed using the bead control sample read counts for the peptides belonging to that bin. After generating a matrix of 115,753 peptide *Z*-scores for 900 MI or 312 EIP samples, we discarded peptides from bacteria, fungi, and allergens from the VirScan library, resulting in 99,460 viral peptides. *Z*-score values were inverse hyperbolic sine-(arcsinh)-transformed in each sample.

Contrarily to log transformation, the arcsinh function is convenient to handle both over-dispersion due to outliers and zero values, which were common in the VirScan *Z*-score data.

Outlier peptides were identified by leveraging replicates through the following process. First, *Z*-score values missing in only one replicate were set to NA in both replicates. Then, outliers in each replicate were defined as *Z*-scores higher than the 99.5% quantile. Next, the absolute difference in *Z*-scores between replicates was calculated for all peptides with an outlier value in at least one replicate. The distribution of absolute differences was bimodal, with the lower peak representing consistent *Z*-scores between replicates and the upper peak representing inconsistent *Z*-scores. The local minimum between the peaks was identified using the optimize function from the *stats* R package, and outliers were defined as all peptides with absolute differences above this minimum. The *Z*-score values of both replicates for all outlier peptides were then set to NA. The rate of missing values was 1.06% in the MI cohort and 1.09% in the EIP cohort. Next, peptides with >50% missing values were removed from the dataset, leaving 98,757 in the MI dataset and 98,697 in the EIP dataset. Duplicated Uniprot entries were removed, leaving 97,975 peptides in the MI dataset and 97,923 in the EIP dataset for the remaining analyses.

Missing values were imputed by first running a PCA on all *Z*-scores using the pca function from the *pcaMethods* package (nPcs = 10, scale = ‘uv’), followed by imputation using the completeObs function from the same package. As individual samples were processed in batches on cell culture plates, samples were batch-corrected using the ComBat^68^ function from the *sva* R package, using plates as the batch variable. The final *Z*-scores were generated by calculating the mean of the two replicates for each individual. A peptide was considered significantly positive if the Z-scores of both replicates were >3.5. The hit variable was defined as 1 if the peptide was positive and 0 otherwise. To generate the list of public peptides, the datasets were filtered on peptides significantly positive in >5% of tested individuals for at least 2 peptides per virus.

### VirScan data processing with AVARDA

Between-species antibody cross-reactivity, unequal representation of viruses in the VirScan library, and viral genome size can make peptide-level data challenging to interpret in some cases. To address these limitations and compare antibody profiles at the virus-species level, we applied the AVARDA algorithm as previously described^34^, using the code available at https://github.com/drmonaco/AVARDA. Briefly, individual VirScan peptides were aligned to each other and to a master library of all viral genetic sequences translated in all reading frames using BLAST. ‘Evidence peptides’ were VirScan peptides that align to the master library with a bit score >80. For each virus, AVARDA calculated a maximally independent set of unrelated peptides that explains the total reactivity towards this virus. A ‘probability of infection’ for each virus was calculated using binomial testing, comparing the ratio of the number of positive evidence peptides to the total number of evidence peptides with the fractional representation of the virus in the VirScan library. Finally, cross-reactivity was evaluated by ranking all viruses based on the probability of infection. Pairs of viruses were then iteratively compared, where shared reactive peptides were assigned to the virus with the most substantial evidence of infection based solely on non-shared peptides. Once all peptides were exclusively assigned to a single virus, a final probability of infection for each sample was calculated using the binomial testing procedure described above. Additionally, a breadth score was calculated, reflecting the total number of positive peptides of independent specificity for each virus.

### Immunoassay-based serological data

Details on the specific antigens and immunoassay methods have been previously described^2^. Blood was collected in EDTA-treated tubes, and the plasma was extracted by centrifugation. Total levels of immunoglobulins IgG, IgM, IgE, and IgA were measured with a turbidimetric test on an Olympus AU400 Chemistry Analyzer. The immunoassay-based serologies were measured for IgG against the following viruses and antigens: CMV (viral lysate), HSV-1 (Glycoprotein G), HSV-2 (Glycoprotein G2), EBV (EBNA-1, VCA p18, EA-D), VZV (Lysate), IAV (Lysate), rubella (Lysate), and measles (Lysate). The data processing steps for the immunoassay-based serology data are described in more detail in ref.^2^. Briefly, the absorbance and emission values collected in each assay are used to call the serostatus for each blood sample. The individual cutoff values used for calling a sample positive or negative are given by the manufacturer and can be found in Table S2 of ref.^2^.

### Luminex-based serological data

MI plasma samples were tested for antibodies to a broad panel of common respiratory pathogens and routine vaccine-preventable diseases using bead-based multiplex assays. A 43-plex assay was developed that included antigens for adenovirus, cytomegalovirus, Epstein-Barr virus, echovirus, enterovirus CoxB3, hepatitis A virus, hepatitis B virus, hepatitis C virus, measles, mumps, rubella, norovirus, respiratory syncytial virus (RSV), rhinovirus, rotavirus, varicella-zoster virus, human papillomavirus, influenza A, human seasonal coronaviruses 229E, NL63, OC43 and HKU1, and SARS-CoV-2. Three antigens for RSV were sourced from The Native Antigen Company (Oxford, UK): RSV A glycoprotein G (RSV-AgG); RSV A lysate (RSV-A); and RSV B lysate (RSV-B). Samples were run at a dilution of 1:100. Plates were read using a Luminex IntelliFlex system, and the median fluorescence intensity was used for analysis. For the Luminex-based serology data, a 5-parameter logistic curve was used to convert median fluorescence intensities to relative antibody units, relative to the standard curve performed on the same plate to account for inter-assay variation.

### Serostatus prediction

We assessed the performance of different methods that predict serostatus from the VirScan data by comparing predicted serostatus to ELISA-based serostatus obtained in the same 900 MI donors. We focused on predicting serostatus for four common viruses for which ELISA data were available: CMV, EBV (EBNA-1 and EA-D), HSV-1, and HSV-2 (Supplementary Note, Table S1). We considered four alternative approaches: (i) the hit-based heuristic method, which assigns seropositivity for a given virus when the number of hits is > 3 or 5 (as in ref.^27^); (ii) the hit-based optimized method, where we searched for the number of positive hits for a given virus that maximizes prediction precision and recall; (iii) the AVARDA-based optimized method, where we searched for the threshold value of the AVARDA breadth score for a given virus maximizes prediction precision and recall, and (iv) an Elastic Net penalized Logistic Regression trained from a subset of the data.

To train the Elastic Net model, we shuffled and split the data into a training set (70% of the data) and a test set (30%) so that the ratio of seropositive to seronegative samples in both sets was the same as in the original data. We only considered VirScan peptide *Z*-scores for the tested virus as features during feature selection. Two complementary approaches were implemented to reduce overfitting: we discarded features with variance lower than a user-specified threshold, defining a first hyper-parameter, and kept the features with univariate association statistics higher than a user-specified percentile, defining a second hyper-parameter. A grid-based approach was used to optimize the two hyper-parameters and the ratio between Elastic Net L1 and L2 penalty, performing a 5-fold cross-validation for each point of the 3-dimensional grid.

We visually inspected learning curves to ensure the absence of overfitting. Processing and modeling were carried out using Python 3.12.2 and the following packages numpy 1.26.4, scipy 1.12.0, pandas 2.2.1 and scikit-learn 1.4.1.post1. All the packages were installed in a conda 24.3.0 environment for reproducibility.

### Kappa-deleting recombination excision circles (KREC) assay

To evaluate if B-cell maturation affects antibody levels, we tested the association between all public peptide *Z*-scores and circulating levels of Kappa-deleting recombination excision circles (KREC), i.e., circular DNA segments generated in B cells during their maturation in bone marrow. KRECs serve as surrogates of new B cell output, as they persist in B cells and get diluted with cell division^69^. KREC quantification was performed as in ref.^70^, with some modifications. Briefly, 1 to 2 µg of whole blood genomic DNA was pre-amplified for 3 minutes at 95°C and then 18 cycles of 95°C for 15 s, 60°C for 30 s and 68°C for 30 s, in a 50 µl reaction containing primers, 200 µM of each dNTP, 2.5 mM MgSO4 and 1.25 unit of Platinum Taq DNA pol High Fidelity (ThermoFisher Scientific, Courtaboeuf, France) in 1× buffer. Forward and reverse primers were TCAGCGCCCATTACGTTTCT and GTGAGGGACACGCAGCC for sjKREC, and CCCGATTAATGCTGCCGTAG and CCTAGGGAGCAGGGAGGCTT for cjKREC, respectively. Probes were CCAGCTCTTACCCTAGAGTTTCTGCACGG (sjKREC) and AGCTGCATTTTTGCCATATCCACTATTTGGAGTA (cjKREC). Columns of 48.48 Dynamic array IFCs (Fluidigm France, Paris, France) were loaded with 5 µl containing 2.25 µl of a 1/2000th dilution of preamplified DNA, 2.5 µl of 2× Takyon Low Rox Probe MM (Eurogentec, Paris, France) and 0.25 µl of sample Loading Reagent and raws with an equal mixture of 2× Assay loading Reagent and 2× Assay Biomark containing only the two primers and the probe specific for each assay and were subjected to a 40 cycles PCR (95°C, 15 s and 60°C, 60 s) in a Biomark HD system (Fluidigm). cjKRECs and sjKRECs were normalized to 150,000 cells using the Albumin gene quantification.

### Genome-wide SNP genotyping

Details about SNP array genotyping of the MI cohort are available elsewhere^45^. Briefly, DNA was extracted from whole blood collected on EDTA using the Nucleon BACC3 genomic DNA extraction kit (catalog #: RPN8512; Cytiva, Massachusetts, USA). The 1,000 MI individuals were genotyped using the HumanOmniExpress-24 BeadChip (Illumina, U.S.), and 966 were also genotyped using the HumanExome-12 BeadChip (Illumina, U.S.). Details about SNP array genotyping of the EIP cohort are available elsewhere^33^. Briefly, PBMCs were isolated from blood collected into EDTA vacutainers, monocytes were removed with CD14+ microbeads, and DNA was isolated from the monocyte-negative fraction using a standard phenol/chloroform protocol, followed by ethanol precipitation. Genotyping was performed in all individuals using the HumanOmni5-Quad BeadChip (Illumina, U.S.) In addition, whole-exome sequencing was performed with the Nextera Rapid Capture Expanded Exome kit.

The genotyping data processing of the MI cohort is described in detail in ref.^45^. After applying quality control filters, the SNP array data sets from the two genotyping platforms were merged. SNPs that were discordant in genotypes or position between the two platforms were removed, yielding a final data set containing 732,341 genotyped SNPs. The data set was then phased using SHAPEIT2^71^ and imputed using IMPUTE v.2^72^, with 1-Mb windows and a buffer region of 1Mb. After imputation, SNPs with an information metric ≤ 0.8, duplicated SNPs, SNPs with a missingness of >5%, and SNPs with a minor allele frequency of ≤5% were removed, generating a final data set of 5,699,237 SNPs. 13 individuals were removed based on relatedness and admixture^45^. Finally, the data set was converted to GRCh38 using the *LiftoverVcf* function from the *GATK* software package^73^.

A more complete description of the genotyping EIP data processing steps can be found in ref^33^. The SNP array genotyping and whole-exome sequencing data were processed separately and merged. For the SNP array data, SNPs were passed through multiple QC filters, and SNPs originating from the sex chromosomes were removed. For the whole-exome sequencing data, reads were processed according to the GATK Best Practices. Discordant variants between the two datasets were removed before merging the SNP array and whole-exome sequencing data sets. After combining the two datasets, the data was phased using SHAPEIT2 and imputed using IMPUTE v.2, with 1-Mb windows and a buffer region of 1 Mb. After imputation and additional QC filtering, 19,619,457 SNPs remained. The data set was converted to GRCh38 using the *LiftoverVcf* function from the *GATK* software package^73^. Finally, four individuals were removed based on relatedness and admixture^33^.

### Whole-genome sequencing

Whole genome sequencing was performed by the Centre National de Recherche en Génomique Humaine (CNRGH), Institut de Biologie François Jacob, Evry, France. After quality control, 1µg of genomic DNA was used to prepare a library using the Illumina TruSeq DNA PCR-Free Library Preparation Kit, according to the manufacturer’s instructions. After normalization and quality control, qualified libraries were sequenced on an Illumina HiSeqX5 platform (Illumina Inc., CA, USA) as paired-end 150 bp reads. One lane of HiSeqX5 flow cell was produced for each sample to reach an average sequencing depth of ∼30× for each sample. FASTQ files were mapped on the human reference genome version hs37d5, using BWA-MEM with default options^74^. BAM file integrity was verified with PicardTools and samtools. Duplicated reads were identified with sambamba^75^. Reads were realigned and recalibrated with GATK^73^ v.4.1.

Sequencing reads mapping to the *HLA*, *IGH*, *IGK,* and *IGL* loci were extracted from the mapped BAM files. Genotypes were called in each individual with HaplotypeCaller in GVCF mode.

Multi-sample genotype calling was performed jointly on combined GVCF files with GATK GenotypeGVCFs. After Variant Quality Score Recalibration (VQSR), variants that passed the tranche sensitivity threshold of 99.0% were selected. Multiallelic sites were split into several biallelic sites with ‘bcftools norm-m-both’ and variants spanning deletions were filtered out. Genotypes were set to missing if the depth of coverage was < 8× or genotype quality < 20. Based on kinship coefficients estimated with KING^76^, ten related individuals and one individual detected as contaminated were excluded. Finally, variants with minor allele frequency (MAF) < 0.05, Hardy-Weinberg equilibrium *P*-value < 10^-10^ (calculated using the *HWExact* function from the *GWASExactHW* R package) or call rate < 0.95 were discarded, resulting in a total of 30,503 common variants near and within immunoglobulin genes.

### Testing association between VirScan *Z*-scores and non-genetic factors

All statistical associations were tested using multiple regression models. In all models, the dependent variable was either an asinh-transformed VirScan Z-score (for a given peptide) or an AVARDA breadth score (for a given virus). The independent variable could be (i) serological measurements based on ELISA, (ii) serological measurements based on Luminex xMAP assays, or (iii) age and sex, continent of birth, and candidate non-genetic factors, including smoking, diet, past diseases, health biomarkers, and anthropometric measures (Table S2). The three variable groups (i), (ii), and (iii) were treated as independent families of tests. Tests within the MI and EIP cohorts were also considered independent. As detailed below, the specific model and complete list of covariates used varied depending on the independent variables being tested.

A linear model was applied using the ‘lm’ R function when the independent variable was continuous or binary. The beta value was used to determine the effect size of the independent variable. When the independent variable was categorical with more than two levels, an ANCOVA model was applied using the ‘aov’ R function. In the association analyses of the MI cohort, age and sex were systematically included as covariates. We also investigated non-linear effects of age by testing an ANOVA model that models age as a factor with five 10-year levels.

In addition, we tested for age×sex interactions by adding an interaction term to the linear model. The only analyzed independent variables for the EIP cohort were age and continent of birth.

When age was used as the variable of interest, the continent of birth was controlled for, and vice versa. As all individuals in the EIP cohort were males, sex was not used as a covariate in these analyses.

To leverage the high resolution of the VirScan peptide library while accounting for between-species antibody cross-reactivity, we first tested the association between non-genetic factors and all public peptide *Z*-scores and then evaluated if AVARDA breadth scores for the tested viruses were associated with the corresponding factors. We considered three scenarios: (i) both the *Z*-scores for several peptides of a given virus and the AVARDA score for the same virus were associated with the candidate factor in the same direction, interpreted as a true association; (ii) the *Z*-scores for several peptides of a given virus were associated with the candidate factor in the same direction, but the AVARDA score for the same virus was not, interpreted as a false association due to cross-reactivity; and (iii) the *Z*-scores for several peptides of a given virus were associated with the candidate factor in opposite directions, but the AVARDA score for the same virus was not associated, interpreted as true associations obscured by opposite epitope-specific effects.

### Testing association between VirScan scores and genetic factors

GWAS was conducted on the asinh-transformed VirScan *Z*-scores or AVARDA breadth scores in the MI cohort. The EIP cohort was used as a replication cohort. The specific covariates used differed between the two cohorts. To correct for population stratification, a principal component analysis was run on all SNPs separately for both cohorts, and the first two principal components were included as covariates. Age was also included as a covariate for both cohorts, and sex was included as a covariate for the MI cohort only. The population of origin was included as an additional binary indicator covariate for the EIP cohort. The GWAS analyses were conducted using the ‘assocRegression’ function from the *GWASTools* R package^77^, using a linear model as the model type and an additive model for the genotype effects. Manhattan plots, locusZoom plots, and tables were all made using the *topr* R package^78^.

### *HLA* allele imputation and association testing

*HLA* allele imputation was done using whole-genome sequencing data of the *HLA* locus (here defined as position 28-35 Mbp in GRCh37), using all variants in the region with MAF ≥ 5%. Imputation was conducted on the Michigan Imputation Server^79^, using the Four-digit Multi-ethnic *HLA* reference panel v2. *HLA* dosages were calculated using plink2^80^. Association testing was conducted similarly to individual SNP analysis but using *HLA* allele dosages instead of SNP genotypes.

### Estimation of the proportion of variance explained

The proportion of variance explained by demographic and genetic factors was estimated for the VirScan *Z*-scores of the 2,608 public peptides in the MI cohort. Genetic factors were the most associated SNPs identified through conditional GWAS, i.e., by testing association with all variants while controlling for hitherto identified lead SNPs. This process was continued until no more SNPs with a *P*-value below genome-wide significance (*P* < 1.31 × 10^-10^) could be identified, leaving a total of 17 SNPs. Age, sex, and smoking were included as demographic factors. The contribution of each of these 20 variables to the variance of each peptide *Z*-score was estimated using the *relaimpo* R package^81^.

### Phylogenetic analyses

All UniProt amino-acid sequences used to build the VirScan peptide library for the RSV protein G were aligned with the msa function from the *msa* package^82^. The 41-aa-long region that was covered by the largest number of UniProt sequences was identified. Based on this shared region, a distance matrix between all Uniprot sequences was computed with the ‘DistanceMatrix’ function from the DECIPHER package^83^, and complete-linkage clustering was used to obtain a phylogenetic tree using the ‘hclust’ R function. Strain annotations were then interpolated for all VirScan peptides using the constructed tree.

---

**Extended Data Fig. 1:**
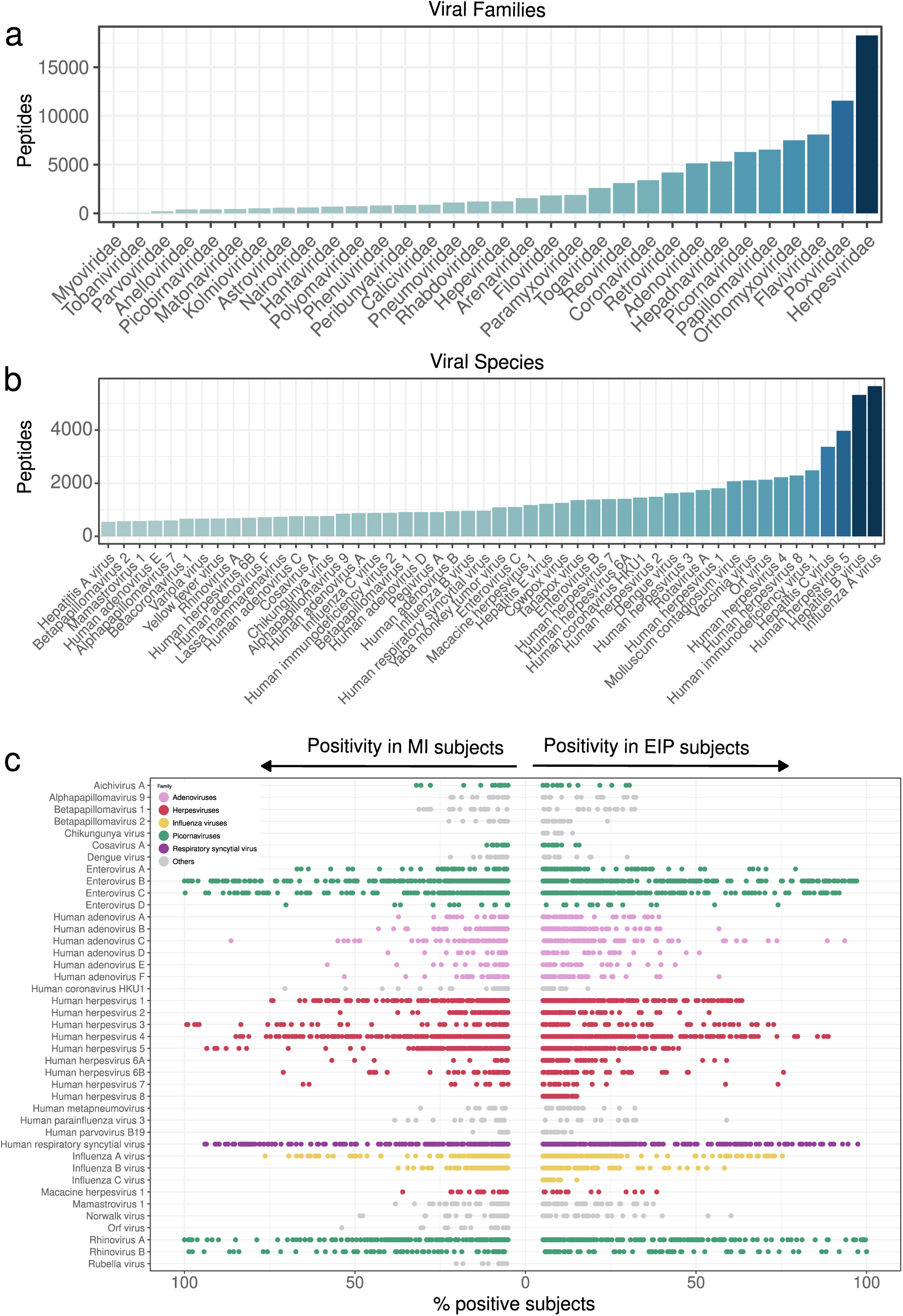
Overview of the viruses targeted by the VirScan assay in the Milieu Intérieur (MI) and EvoImmunoPop (EIP) cohorts. **a,b**, Number of peptides in the VirScan PhIP-seq library, separated by viral family (**a**) and viruses (**b**). Only the 50 most covered viruses are shown. **c**, Percentage of MI (left) and EIP (right) individuals positive for 2,608 public peptides, separated by virus. Each point indicates a viral peptide, colored according to its viral family. Only viruses with at least 10 peptides showing an enrichment of >5% are included.

**Extended Data Fig. 2:**
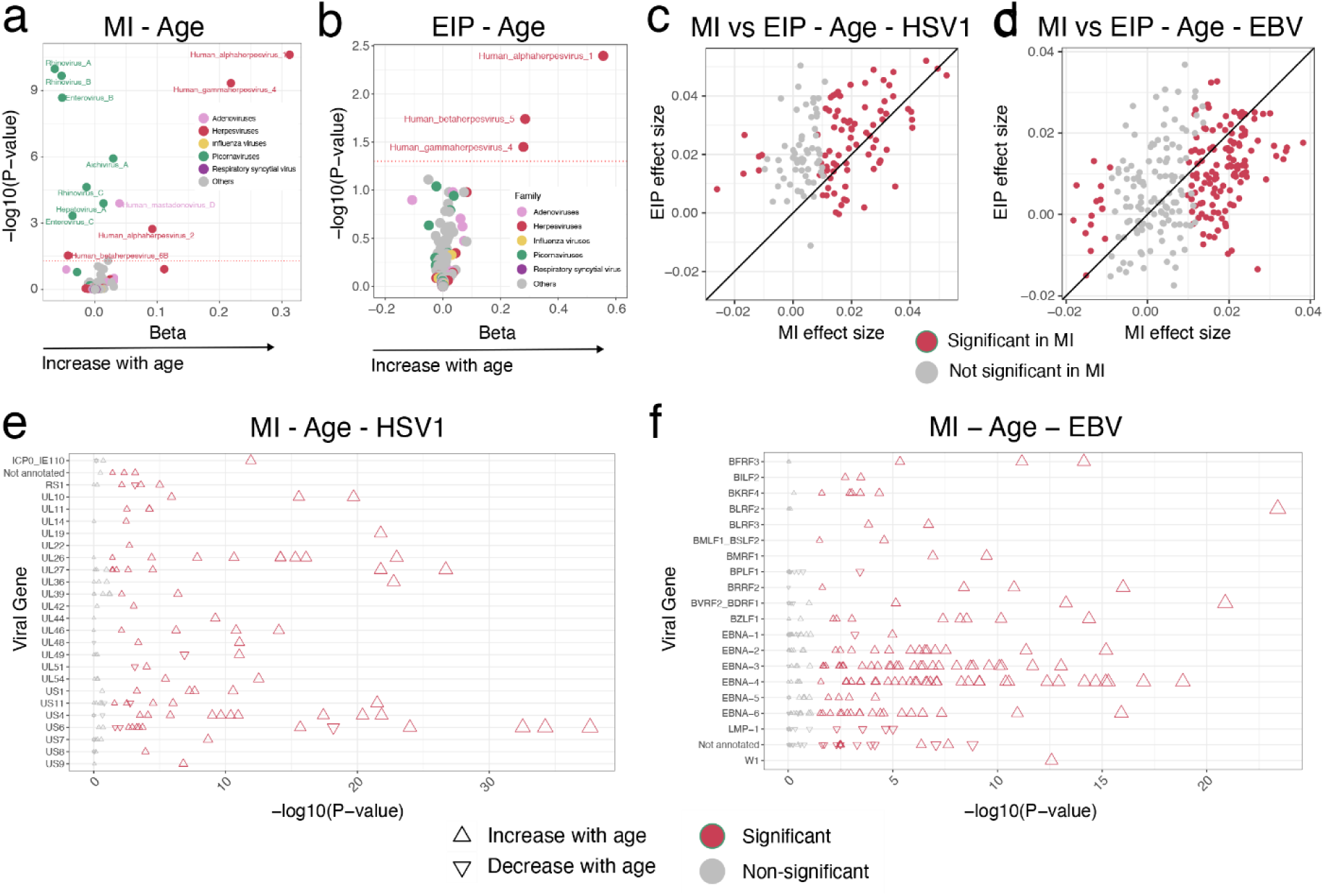
Additional age differences in the antiviral antibody repertoire. **a,b**, – log10(adjusted *P*-values) against effect sizes for associations between the AVARDA breadth score and age in the MI (**a**) and EIP (**b**) cohorts. Each point indicates a virus, colored according its viral family. **c,d**, Effect sizes for the associations between age and HSV-1 (**c**) and EBV (**d**) peptide *Z*-scores in the MI and EIP cohorts. **e,f**, –log10(adjusted *P*-values) of associations between age and HSV-1 (**e**) and EBV (**f**) peptide *Z*-scores, separated by viral protein, in the MI cohort. The significance and direction of associations are indicated by color and by the direction of triangles, respectively.

**Extended Data Fig. 3.**
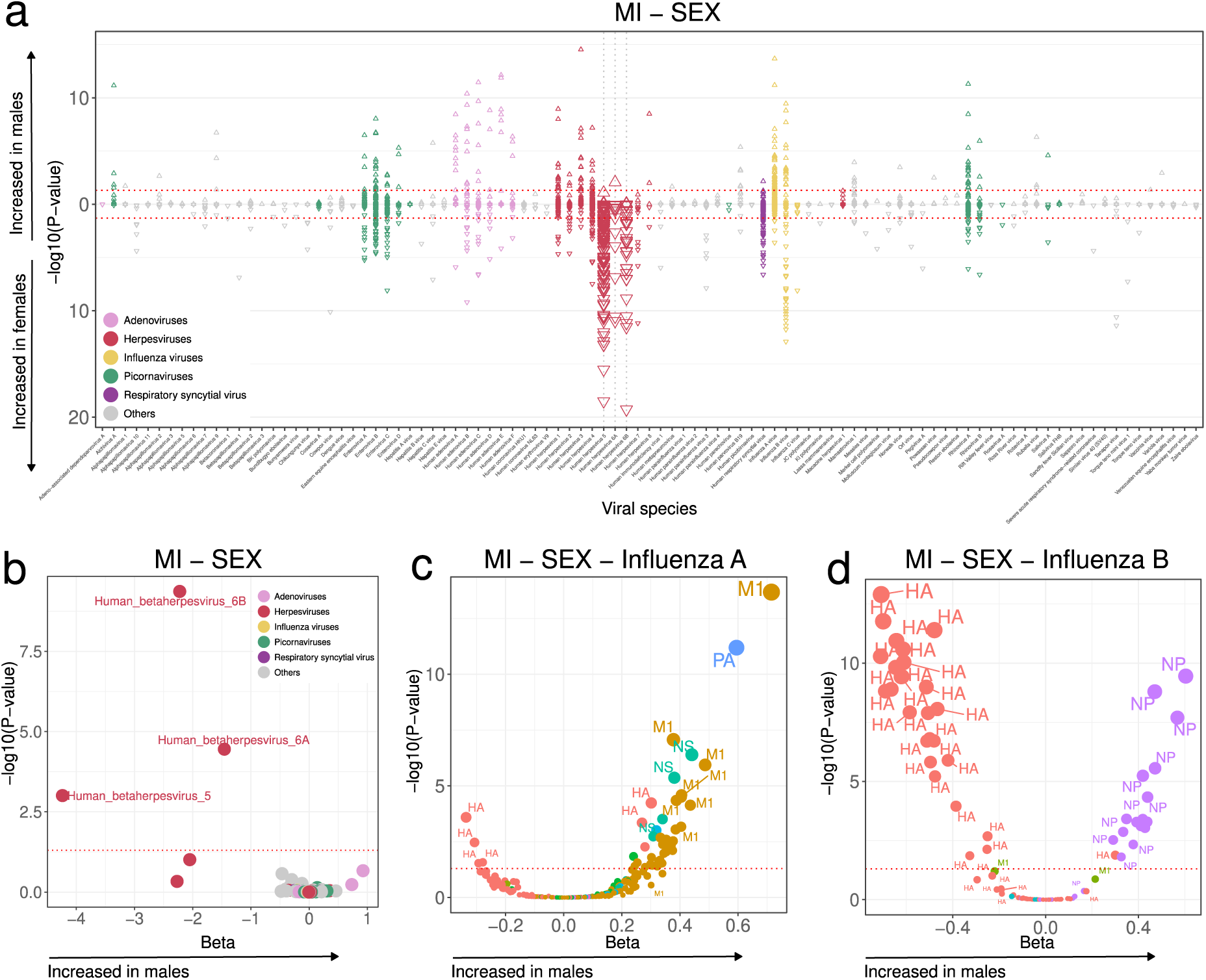
Sex differences in the antiviral antibody repertoire. **a**, –log10(adjusted *P*-values) and direction of associations between all public peptide *Z*-scores and sex in the MI cohort. **b**, –log10(adjusted *P*-values) against effect sizes for associations between the AVARDA breadth score and sex in the MI cohort. Each point indicates a virus, colored according to its viral family. **c,d**, –log10(adjusted *P*-values) against effect sizes for associations between IAV (**c**) and IBV (**d**) peptide *Z*-scores and sex in the MI cohort, colored according to the viral protein.

**Extended Data Fig. 4.**
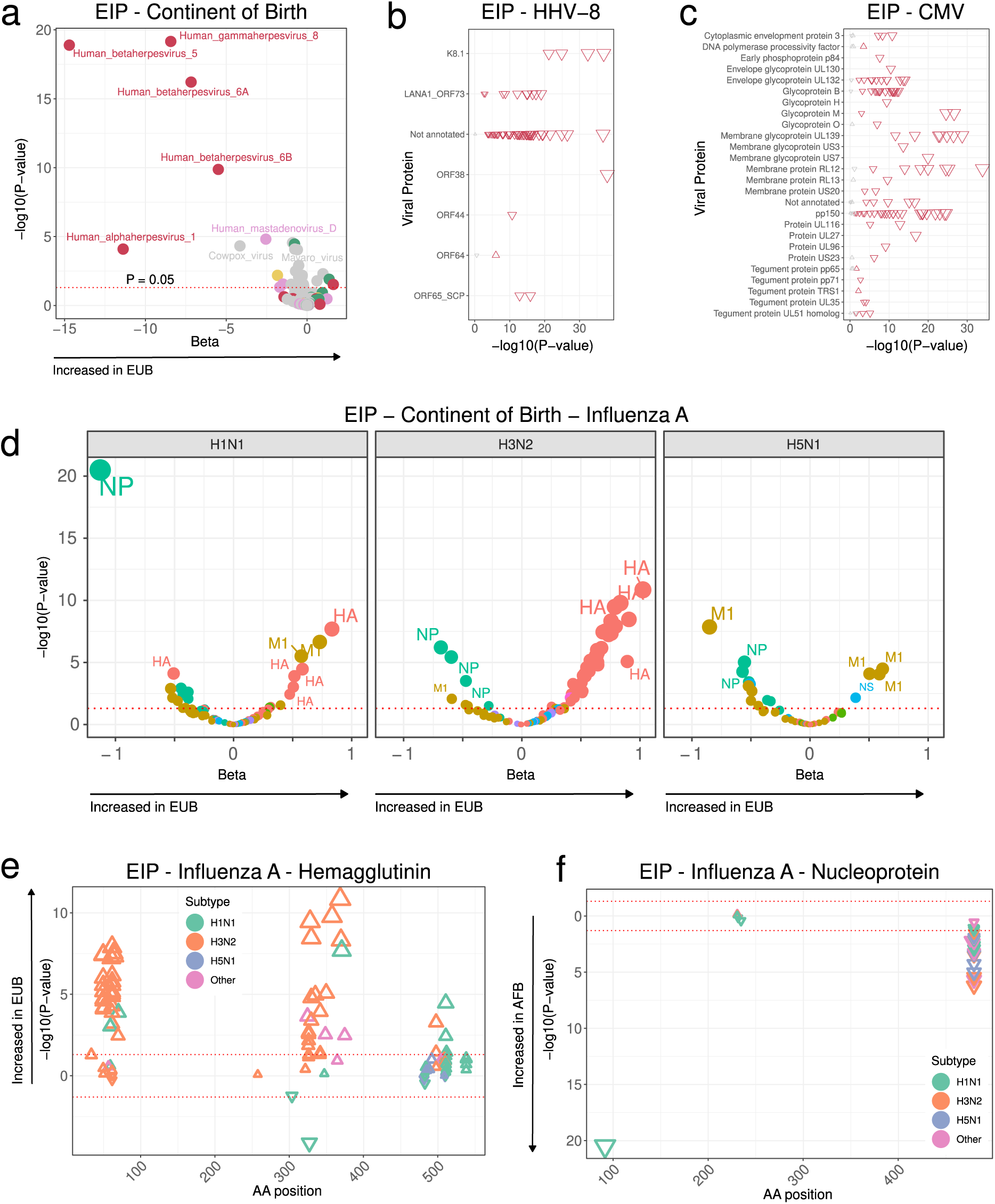
Additional population differences in the antiviral antibody repertoire. **a**, –log10(adjusted *P*-values) against effect sizes for associations between the AVARDA breadth score and continent of birth in the EIP cohort. Each point indicates a virus, colored according to its viral family. **b,c,** –log10(adjusted *P*-values) of associations between continent of birth and HHV-8 (**b**) and CMV (**c**) peptide *Z*-scores, separated by viral protein, in the EIP cohort. The significance and direction of associations are indicated by color and by the direction of triangles, respectively. **d,** –log10(adjusted *P*-values) against effect sizes for associations between IAV peptide *Z*-scores and continent of birth in the EIP cohort, faceted by main IAV subtypes. Colors indicate the viral protein. **e,f,** Amino-acid positions of the midpoint of HA (**e**) and NP (**f**) peptides associated with continent of birth within the full IAV proteins for the EIP cohort. The significance and direction of associations with age are indicated on the y-axis and by the direction of triangles, respectively. The triangle color indicates the IAV subtype.

**Extended Data Fig. 5.**
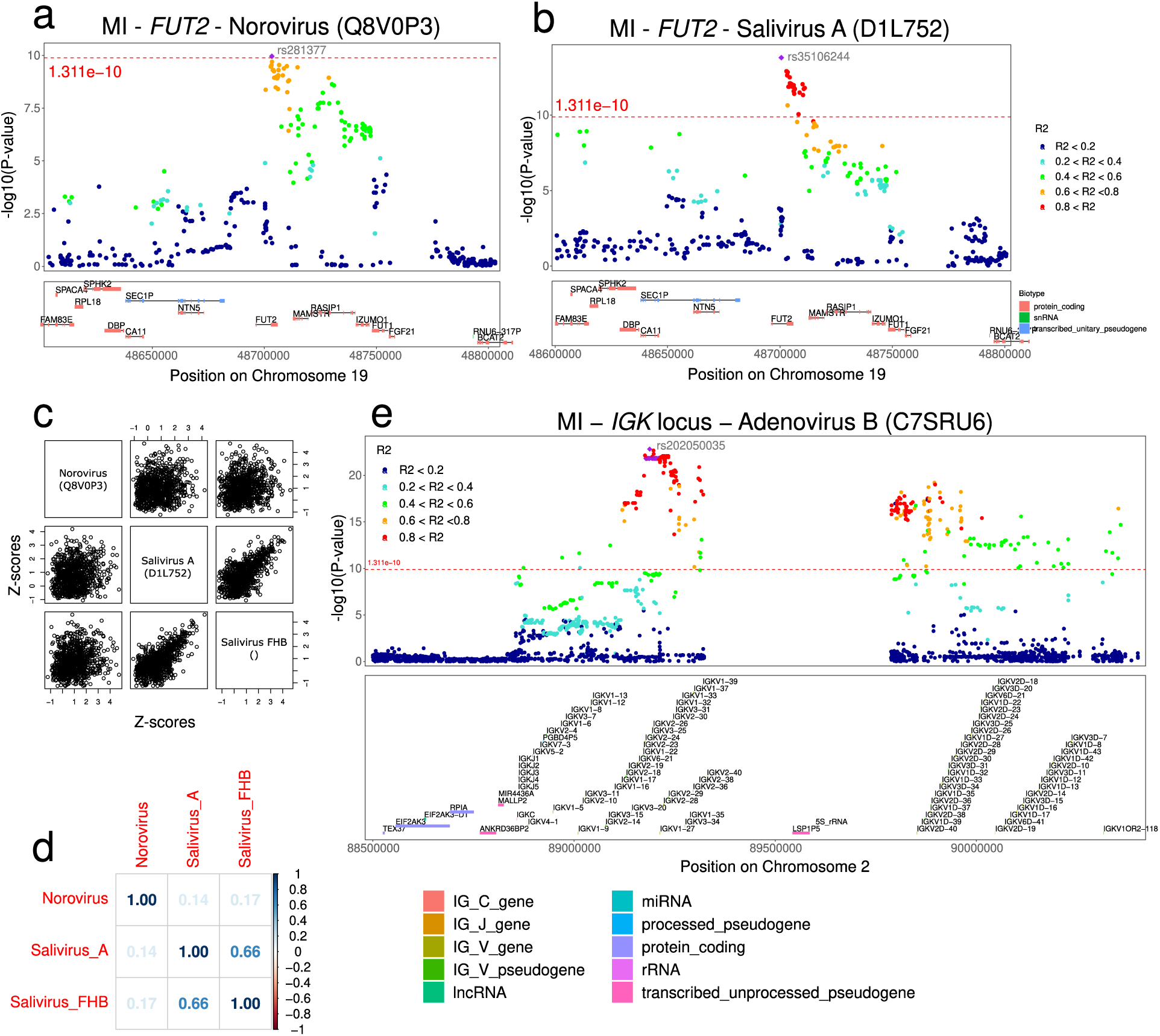
Association of genetic variation in the *FUT2* and *IGK* loci with the antiviral antibody repertoire. **a,b,** LocusZoom plots showing associations between the *FUT2* locus and antibody reactivity against (**a**) norovirus (UniProt ID: Q8V0P3) and (**b**) salivirus A (UniProt ID: D1L752). **c,d**, Scatter plots (**c**) and correlation matrix (**d**) for the three norovirus and salivirus peptide *Z*-scores most significantly associated with *FUT2* variants. **e**, LocusZoom plot showing associations between the *IGH* locus and antibody reactivity against adenovirus B (UniProt ID: C7SRU6).

**Extended Data Fig. 6.**
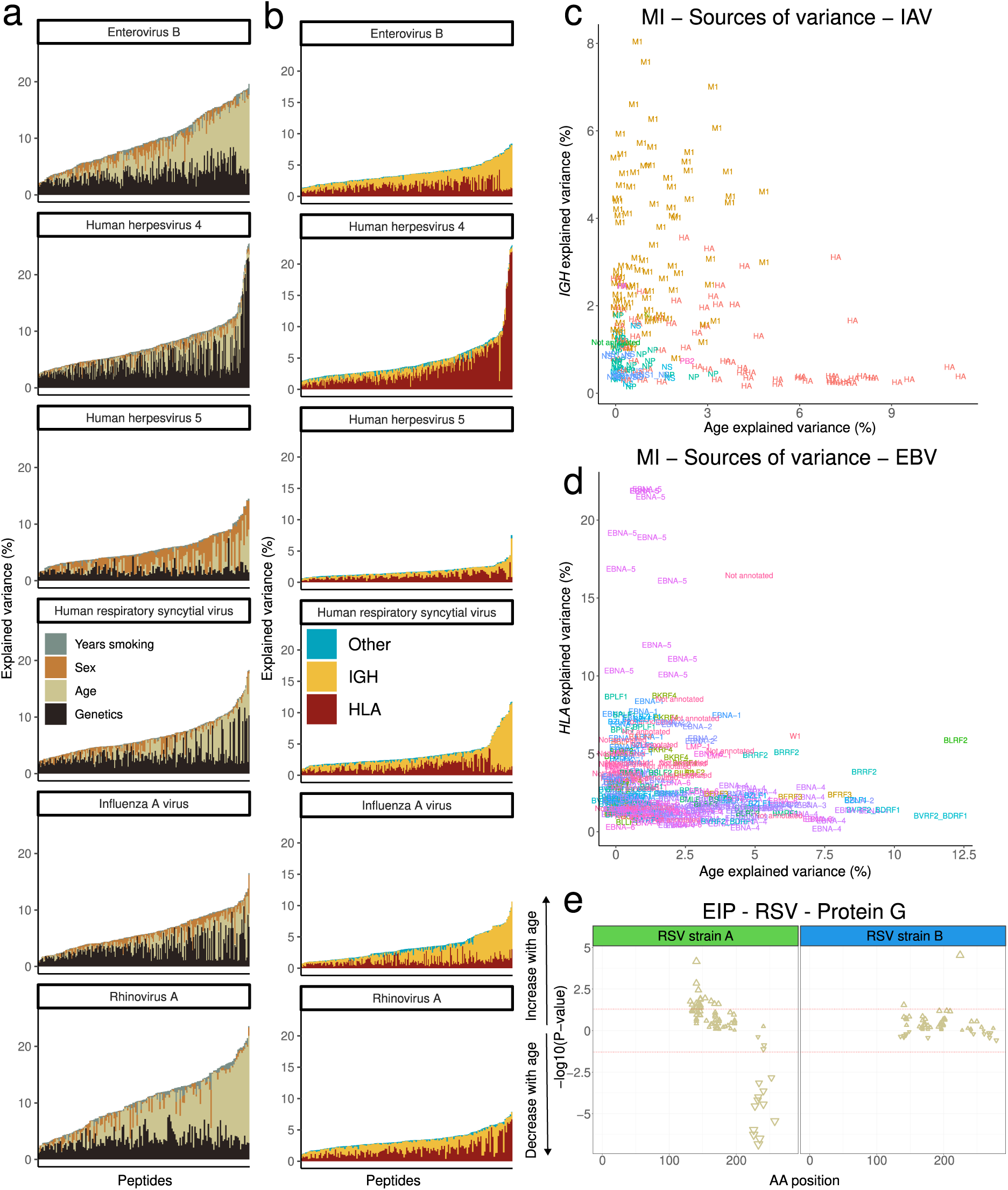
Variance in the antiviral antibody repertoire explained by individual factors. **a**, Proportion of variance explained by age, sex, smoking, and genetics for antibody reactivity against public peptides from the six viruses with the largest number of public peptides in the MI cohort. Peptides are sorted by total variance explained. **b**, Proportion of variance explained by genetic factors for antibody reactivity against public peptides from the six viruses with the largest number of public peptides in the MI cohort. Genetic variance is separated by genetic variation in the *HLA* and *IGH* loci and variation external to these loci. Peptides are sorted by total variance explained. **c,d,** Variance explained by age and *IGH* genetic variation for IAV peptides (**c**) and age and *HLA* genetic variation for EBV peptides (**d**) in the MI cohort, colored according to protein. **e**, Amino-acid positions of the midpoint of protein G peptides associated with continent of birth within the full RSV protein G for the EIP cohort. The significance and direction of associations are indicated on the y-axis and by the direction of triangles, respectively.

