## Supplementary Note for "Demographic and genetic factors shape the epitope specificity of the human antibody repertoire against viruses"

### Validation of the VirScan PhIP-seq results using ELISA- and Luminex-based assays

To validate the results collected through the PhIP-seq approach, we measured antibody reactivity against 55 antigens from 21 viruses using either ELISA ($n$ = 12) or Luminex xMAP ($n$ = 43) assays in the same individuals from the MI cohort[1] (Methods). We compared PhIP-seq-based estimates of antibody reactivity to ELISA or Luminex serological results by first testing the association with 132 AVARDA breadth scores. We found that AVARDA breadth scores were specifically associated with the ELISA serostatus for the corresponding virus for 67% (8/12) of assays (Supplementary Fig. 1a), including herpesviruses such as CMV ($P_{adj}$ = $9.24 \times 10^{-160}$) and EBV ($P_{adj}$ = $2.72 \times 10^{-19}$) (Supplementary Fig. 1b-c), as well as the rubella ($P_{adj}$ = $4.43 \times 10^{-2}$) (Supplementary Fig. 1d) and varicella zoster ($P_{adj}$ = $3.73 \times 10^{-2}$) viruses. For the remaining assays, including IAV ($P_{adj}$ = 0.38) and HBV ($P_{adj}$ = 0.63), ELISA serostatus was associated with none of the AVARDA breadth scores (Supplementary Fig. 1e-f). Similarly, of the 43 Luminex assays, 62.8% (27/43) were significantly associated with an AVARDA breadth score, whereas 16 were not. Of the 27 significant associations, 77.8% (21/27) were found with the cognate virus (Supplementary Fig. 1g). Akin to the ELISA assays, the viruses most significantly associated with Luminex-based serology included CMV ($P_{adj}$ = $1.50 \times 10^{-171}$) and EBV ($P_{adj}$ = $1.48 \times 10^{-42}$) (Supplementary Fig. 1h-i). Notably, the associations between the VirScan AVARDA scores and the Luminex-based serologies assay were stronger than those with ELISA-based serologies for several viruses, including measles, mumps, rubella, and influenza (Supplementary Fig. 1).

Several inherent methodological differences could explain the discrepancies observed between the PhIP-seq-based AVARDA breadth scores and the ELISA and Luminex assays: (i) the latter assays may target epitopes that are not the most immunoreactive, (ii) the AVARDA algorithm may discard the most immunoreactive peptides, (iii) the AVARDA algorithm may select cross-reactive peptides, and (iv) the VirScan assay may poorly measure reactivity against a specific epitope, due to the use of linear peptides or measurement errors. To evaluate these scenarios, we then tested the association between VirScan $Z$-scores for 2,608 public peptides and ELISA and Luminex serologies in the MI cohort (Methods). Associations between peptide $Z$-scores and serostatus were most significant for the cognate virus for 75% (9/12) of ELISA assays and 58% (25/43) of Luminex assays (Supplementary Fig. 2a-b). Herpesviruses were again strongly associated between serological data sets. For example, 137 out of 180 VirScan peptides significantly associated with ELISA CMV serostatus originated from CMV ($P_{adj} = 7.84 \times 10^{-309}$). Similarly, 137 out of 169 VirScan peptides significantly associated with Luminex CMV serostatus originated from CMV ($P_{adj} = 2.02 \times 10^{-227}$) (Supplementary Fig. 2c-d). These results also revealed a few cases of cross-reactivity for the peptide-level PhIP-seq data. For example, CMV serostatus was significantly associated with several peptides from non-CMV viruses, including Enterovirus B ($P_{adj} = 8.35 \times 10^{-47}$), Tanapox ($P_{adj} = 3.83 \times 10^{-45}$), and Ebola ($P_{adj} = 4.34 \times 10^{-39}$) viruses.

Interestingly, there were cases where peptide *Z*-scores were strongly and specifically associated with ELISA- or Luminex-based serostatus, but the AVARDA score was not. For example, peptide *Z*-scores originating from IAV were significantly associated with both ELISA- (71 significant peptides, $\min(P_{adj}) = 6.01 \times 10^{-7}$) and Luminex-based (137 significant peptides, $\min(P_{adj}) = 1.61 \times 10^{-26}$) IAV serostatuses (Supplementary Fig. 2e-f), whereas the AVARDA score for IAV was not (Supplementary Fig. 1e). This indicates that the peptide aggregation performed by the AVARDA algorithm can lead to false negatives and that increased resolution can be achieved by analyzing individual peptides.

VirScan also identified distinct reactivity patterns against different antigens from the same virus. For example, we found strong associations between ELISA-based EBV serostatus for EA, VCA, and EBNA antigens and the corresponding VirScan peptides (Supplementary Fig. 2g). Additionally, VirScan confirmed the known discordance between EBV serological markers[2], including a small number of VCA-positive, EBNA-negative individuals, and an even smaller group of VCA-negative, EBNA-positive individuals. Discordances among Luminex-based serologies were also apparent when comparing antibody titers against the nucleocapsid protein and the spike protein in some coronaviruses (Supplementary Fig. 2h). Collectively, these analyses show that VirScan PhIP-seq is overall specific and provides high resolution and sensitivity.

**Serostatus prediction by machine learning outperforms heuristic methods**

Defining serostatus from serological data, including PhIP-seq data, is a long-standing challenge because antibody titers often follow a continuous distribution and may target different antigens within the same virus. Previous studies using VirScan have defined seropositivity based on an arbitrary threshold of three to five positive peptides (or ‘hits’) without assessing the performance of this heuristic approach[3]. We thus leveraged the MI data to assess the prediction performance of three alternative approaches: (i) the hit-based heuristic method (‘Hit-H method’), which assigns seropositivity for a given virus when the number of hits is > 3 or 5 (as in ref.[3]); (ii) the hit-based optimized method ('Hit-O method'), where we searched for the number of positive hits for a given virus that maximizes prediction precision and recall; and (iii) the AVARDA-based optimized method ('AVARDA-O method'), where we searched for the threshold value of the AVARDA breadth score for a given virus maximizes prediction precision and recall. Given the strong and specific associations observed between VirScan peptide *Z*-scores and gold-standard ELISA serostatuses (Supplementary Fig. 1 & 2), we also trained a predictive model of serostatus using logistic regression with Elastic Net penalty ('EN method'). Prediction performance was estimated by out-of-sample 5-fold cross-validation, keeping 30% of the MI data as the test set. As our primary objective was to provide a proof-of-concept that machine learning outperforms other approaches, we focused on predicting serostatus for four common viruses for which ELISA data were available: CMV, EBV (EA and EBNA), HSV-1, and HSV-2.

We found that the EN method outperformed the alternative approaches in almost all cases. For CMV, the machine-learning model showed a precision and recall of 97.8% and 97.8%, respectively (Table S1). We estimated a similar performance for the Hit-O method (95.7% and 98.9%), which uses 14 hits, whereas the Hit-H method, using 5 hits, yielded 36.6% precision and 100% sensitivity. The AVARDA-O method showed lower performance than the EN and Hit-O approaches. For HSV-1, EN model precision and recall were 97.3% and 96.8%, versus 87.5% and 100% for the Hit-H approach. The Hit-O method slightly outperformed EN in terms of both precision and recall (Table S1). Differences in performance were more evident for HSV-2, with 93.9% precision and 92.4% recall for EN, while the alternative methods all showed <70% precision. Finally, EBV seropositivity (EBNA and EA epitopes) was predicted by EN with higher accuracy than any other method. Overall, these results indicate that the heuristic approach used in previous studies has relatively poor performance, particularly when the number of seropositive and seronegative samples is unbalanced. In contrast, predicting gold-standard serostatus using a machine-learning method trained on the VirScan data yields highly accurate results.

**Socio-economic status and health biomarkers are weakly associated with the antibody repertoire**

To identify demographic factors affecting the antiviral antibody repertoire, we searched for associations between VirScan peptide *Z*-scores and a curated list of 108 variables assessing socio-economic status (SES), health-related habits, medical history, and health biomarkers collected in the MI cohort (Table S2), while controlling for age, sex, and genetic structure

(Methods). In addition to the strong effects of smoking behavior described (Fig. 4b-e), we found that 65 variables were significantly associated with antibody reactivity against at least one viral peptide ($P_{adj} < 0.05$; Fig. 4a).

Among the strongest associations, we found increased antibodies against enteroviruses in individuals who live with children ($P_{adj} = 0.0014$; Fig. 4a), who recently experienced a depressive episode ($P_{adj} = 2.45 \times 10^{-5}$), and whose highest diploma was high school ($P_{adj} = 0.0062$). High educational attainment was also associated with lower antibodies against HSV-1 ($P_{adj} = 0.0018$), confirming previous seroprevalence surveys[4]. Regarding health biomarkers, we detected a relatively strong, negative association between total protein levels and anti-RSV antibodies ($P_{adj} = 2.61 \times 10^{-5}$; Fig. 4a). Previous work has suggested that RSV infection induces a reduction in proteins, particularly surfactant proteins, which may contribute to RSV pathogenesis in the lung[5]. Additionally, we found that bilirubin levels were associated with lower antibody levels against HHV6A/B ($P_{adj} = 5.34 \times 10^{-5}$) and rhinoviruses ($P_{adj} = 8.82 \times 10^{-4}$), which  remained significant after adjusting for smoking status ($P_{adj} = 2.50 \times 10^{-3}$). Lastly, mean corpuscular hemoglobin concentration was associated with anti-HSV-1 antibodies ($P_{adj} = 1.36 \times 10^{-4}$). Together, these findings suggest that antibody reactivity against common viruses is weakly related to socio-economic status and several health biomarkers in a healthy population.

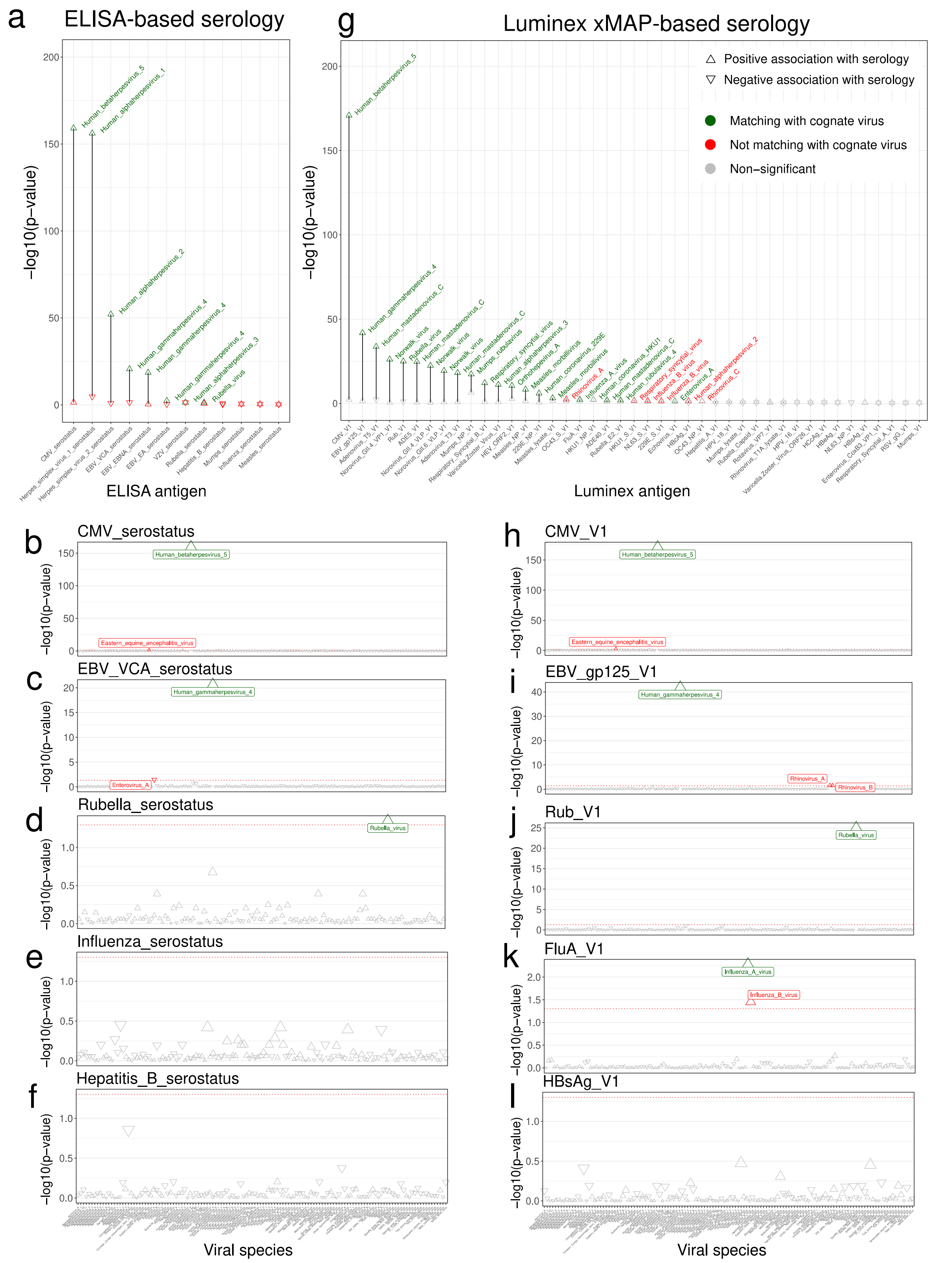


**Supplementary Figure 1: Validation of AVARDA scores by comparisons to ELISA- and Luminex-based serostatuses in the MI cohort. a,g,** –log$_{10}$($P$-values) for the association between serology determined by (**a**) ELISA or (**g**) Luminex xMAP and the AVARDA breadth scores. Serology variables are plotted on the x-axis. The top two AVARDA associations are connected by a black vertical line. Significant associations (FDR < 0.05) are colored in green or red if the association is or is not for the cognate virus, respectively. Non-significant associations are colored gray. **b-f**, –$\log_{10}$(*P*-values) for the association between the AVARDA breadth scores and ELISA-based serostatus for (**b**) CMV, (**c**) EBV VCA antigen, (**d**) rubella virus, (**e**) IAV, and (**f**) hepatitis B. **h-l**, –$\log_{10}$(*P*-values)for the association between the AVARDA breadth scores and Luminex-based serology for (**h**) CMV, (**i**) EBV VCA antigen, (**j**) rubella virus, (**k**) IAV, and (**l**) hepatitis B. Significant associations (FDR < 0.05) are colored in green or red if the association is or is not for the cognate virus, respectively. Non-significant associations are colored gray.

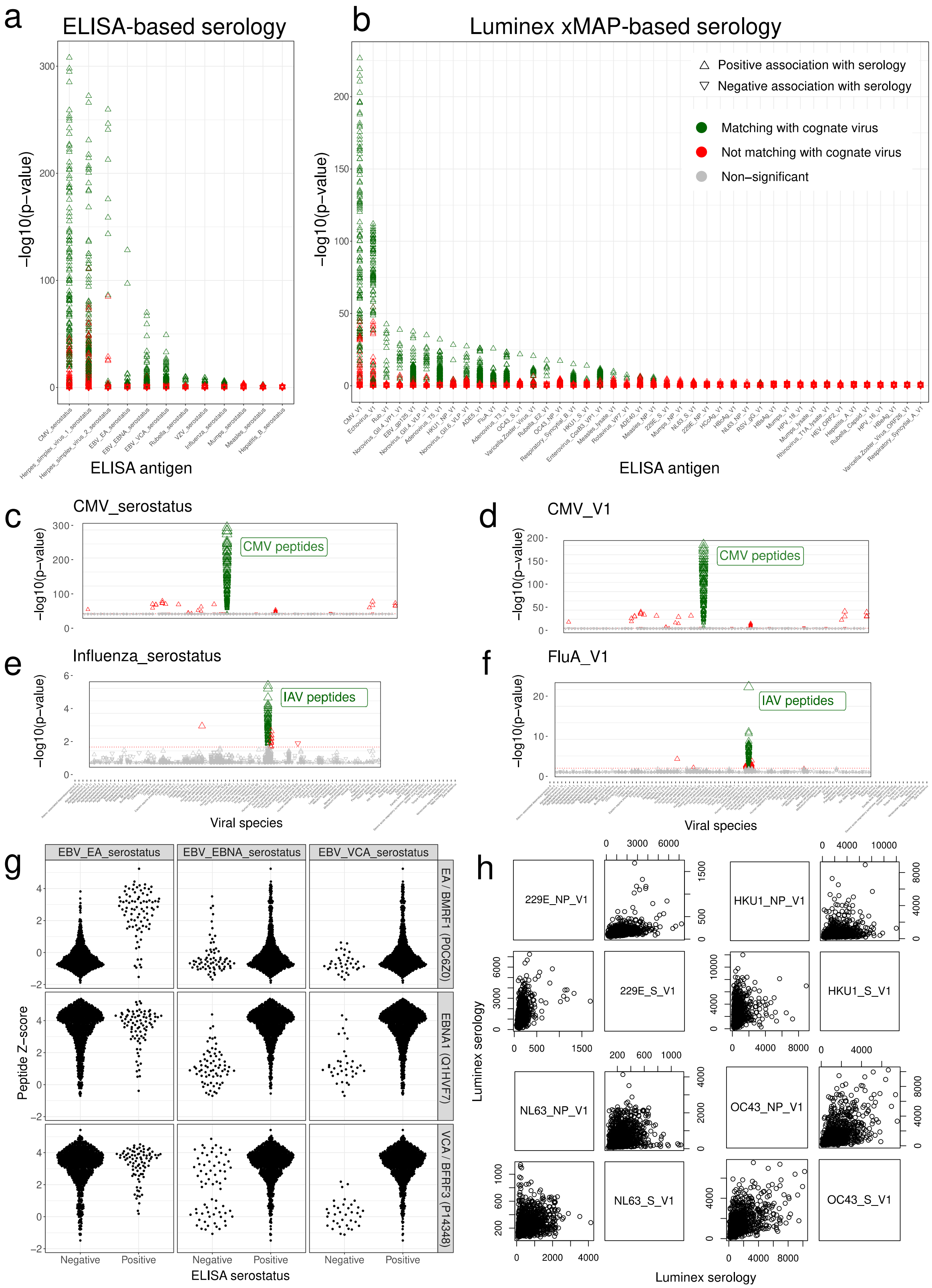


**Supplementary Figure 2: Validation of VirScan Z-scores by comparisons to ELISA- and Luminex-based serostatuses in the MI cohort. a,b**, –log$_{10}$($P$-values) for the association between the VirScan Z-scores and serology determined by (**a**) ELISA or (**b**) Luminex xMAP. Serology variables are plotted on the x-axis, and for each serology variable, the –log$_{10}$($P$-

values) for the association with all 2,608 public VirScan peptides are shown on the y-axis. Significant associations (FDR < 0.05) are colored in green or red if the association is or is not for the cognate virus, respectively. Non-significant associations are colored gray. **c,e**, $-\log_{10}$(*P*-values) for the association between the VirScan *Z*-scores and ELISA-determined serostatus for (**c**) CMV and (**e**) IAV. Significant associations (FDR < 0.05) are colored in green or red if the association is or is not for the cognate virus, respectively. Non-significant associations are colored gray. **d,f**, $-\log_{10}$(*P*-values) for the association between the VirScan *Z*-scores and Luminex-determined serology for (**d**) CMV and (**f**) IAV. Significant associations (FDR < 0.05) are colored in green or red if the association is or is not for the cognate virus, respectively. Non-significant associations are colored gray. **g,** VirScan *Z*-score distributions for the three peptides most significantly associated with ELISA-based serostatus for the EBV antigens EA, VCA, and EBNA. ELISA-based serostatus for each EBV antigen is shown on the x-axis. **h**, Luminex-based serologies for the spike and nucleocapsid proteins shown for the four coronaviruses 229E, HKU1, NL63, and OC43.
